# Trans-ancestral Genome Wide Association Study of Sporadic and Recurrent Miscarriage

**DOI:** 10.1101/2024.03.20.24304624

**Authors:** Alexandra Reynoso, Priyanka Nandakumar, Jingchunzi Shi, Jessica Bielenberg, 23andMe Research Team, Michael V. Holmes, Stella Aslibekyan

**Author notes:** Corresponding authors: Alexandra Reynoso, MSc Stella Aslibekyan, PhD. Joint senior authors (Michael V. Holmes, Stella Aslibekyan).

## Abstract

Miscarriage is a common adverse pregnancy outcome, impacting approximately 15% of pregnancies. Herein, we present results of the largest trans-ancestral genome wide association study for miscarriage to date, based on 334,593 cases of sporadic, and 52,087 cases of recurrent miscarriage in the 23andMe, Inc. Research Cohort. We identified 10 novel genome-wide significant associations for sporadic miscarriage, and one for recurrent miscarriage. These loci mapped to genes with roles in neural development and telomere length, and to developmental disorders including autism spectrum disorder. Three variants, with similar directionality and magnitude of effect, replicated in a previously published GWAS. Using Mendelian randomization and triangulation, robust evidence was found for smoking causally increasing the risk of sporadic (genetic liability to ever vs never smoking: OR 1.13; 95%CI: 1.11-1.15; P=2.61e-42) and recurrent (OR 1.25; 95%CI: 1.21-1.30; P=5.47e-34) miscarriage, with moderate, yet triangulating, evidence identified for a potential etiological role of caffeine consumption.

## Background

Miscarriage is defined as the spontaneous loss of a clinically established pregnancy before 24 weeks of gestation^1^. It is the most common pregnancy complication and affects around 15% of clinically confirmed pregnancies ^2^. Roughly one quarter of all people with a uterus will undergo at least one miscarriage in their lives ^1^.

Miscarriage is considered to arise from the interaction between genetic, environmental, and lifestyle factors, with the risk increasing with parental age ^3,4^. The genetic architecture behind predisposition to miscarriage includes embryonic aneuploidy, chromosomal abnormalities in the parent, and parental genetic polymorphisms. Previous studies have demonstrated the importance of functional genetic knockouts in the etiology of miscarriage ^3,5^, and common genetic polymorphisms related to inflammation and apoptosis have been implicated in risk of recurrent miscarriage ^6^.

Prior genome-wide association studies have typically focused on recurrent pregnancy loss and/or were based on relatively small sample sizes ^3,7^. We conducted a large-scale genome wide association study across three ancestral groups to shed light on the genetic architecture predisposing risk of miscarriage and used Mendelian randomization to explore the potential causal roles of smoking and caffeine.

## Results

### Descriptive analysis

A total of 910,889 participants were included in the descriptive analysis of the study: 340,751 sporadic miscarriage cases, including 50,861 recurrent miscarriage cases nested within sporadic miscarriage cases, and 570,138 controls (Supplementary Figure 1). Among study participants, 86% of individuals were of European ancestry, 11% were of Latinx ancestry, and 4% were of African American ancestry (Table 1). Endometriosis, ovarian cysts, uterine fibroids, or PCOS were more common among individuals with sporadic or recurrent miscarriage. Similarly, a history of ever smoking was more prevalent among both recurrent and sporadic miscarriage cases compared to controls. Daily caffeine consumption was lower among controls (155.5 mg) compared to either recurrent (159.64 mg) or sporadic miscarriage cases (159 mg). Individuals who experienced recurrent miscarriage had on average 5 pregnancies, compared to sporadic miscarriage cases (mean of 4 pregnancies) and controls (mean of 2 pregnancies).

**Table 1:** Demographic, lifestyle, and health characteristics by miscarriage variable (sporadic or recurrent) vs. controls. The survey that collected data was targeted towards participants with self-reported infertility or subfertility, hence the proportions may not be representative of the underlying cohort. Survey data were collected at the of analysis, therefore, reported diseases and/or behaviors may have been different compared to time of miscarriage.

|  |  | Miscarriage Cases |  | Controls<br>(n =570,138) |
| --- | --- | --- | --- | --- |
|  |  | Recurrent<br>(n =50,861) | Sporadic<br>(n =340,751) |  |
| **Age (determined at the time of analysis), mean (SD), years** |  | 55.47 (13.36) | 56.03 (14.02) | 56.77 (14.53) |
| **Ancestry Group** |  |  |  |  |
|  | **European** | 43,446 (85.42%) | 290,391 (85.22%) | 491,102 (86.14%) |
|  | **Latinx** | 5,545 (10.90%) | 37,800 (11.09%) | 60,646 (10.64%) |
|  | **African American** | 1,870 (3.68%) | 12,560 (3.69%) | 18,390 (3.23%) |
| **Education** |  |  |  |  |
|  | **High School Diploma and below** | 5,354 (10.53%) | 37,736 (11.07%) | 64,954 (11.39%) |
|  | **Some College, Associate's Degree, Bachelor's Degree** | 31,072 (61.09%) | 205,413 (60.28%) | 336,792 (59.07%) |
|  | **Master's Degree and above** | 10,312 (20.27%) | 68,026 (19.96%) | 118,079 (20.71%) |
| **Body mass index, mean (SD), kg/m<sup>2</sup>** |  | 28.98 (7.24) | 28.56 (6.98) | 28.22 (6.79) |
| **Ever tobacco smoker, N (%)** |  | 23,681 (46.56%) | 150,303 (44.11%) | 234,465 (41.12%) |
| **Daily alcohol consumption, mean (SD), servings** |  | 0.72 (1.21) | 0.78 (1.21) | 0.72 (1.23) |
| **Current daily caffeine consumption, median (IQR), mg** |  | 159.64 (307.36) | 159 (291.39) | 155.5 (285) |
| **Age of menarche, mean (SD), years** |  | 12.27 (1.75) | 12.34 (1.66) | 12.41 (1.58) |
| **Number of biological children, mean (SD)** |  | 2.14 (1.72) | 2.06 (1.42) | 1.97 (1.72) |
| **Number of pregnancies, mean (SD)** |  | 5.05 (1.21) | 3.63 (1.49) | 2.26 (1.11) |
| **Age of first pregnancy, mean (SD), years** |  | 22.48 (5.90) | 23.08 (5.81) | 23.79 (5.51) |
| **Parental Reproductive Outcomes** |  |  |  |  |
|  | **PCOS, N (%)** | 7,243 (14.24%) | 34,764 (10.20%) | 41,477 (7.27%) |
|  | **Preeclampsia, N (%)** | 6,462 (12.71%) | 36,926 (10.84%) | 51,818 (9.09%) |
|  | **Hyperandrogenism, N (%)** | 1,104 (2.17%) | 5,436 (1.60%) | 7,041 (1.23%) |
|  | **Endometriosis, N (%)** | 11,185 (21.99%) | 57,974 (17.01%) | 74,383 (13.05%) |
|  | **Ovarian cyst, N (%)** | 21,695 (42.66%) | 122,215 (35.87%) | 166,090 (29.13%) |
|  | **Uterine Fibroids, N (%)** | 10,360 (20.37%) | 60,850 (17.86%) | 88,656 (15.55%) |

### GWAS

A trans-ancestral GWAS meta-analysis of sporadic miscarriage was conducted using 334,593 cases and 505,008 controls, after filtering based on relatedness. GWAS identified 10 significant loci (Figure 1, Supplementary Figure 3, Table 2, Supplementary Table 7, Supplementary Table 8). The top hit, rs13107325 (P=1.6e-10), is a missense variant in *SLC39A8*, which encodes the primary transporter of toxic cation cadmium found in tobacco.^8^ The credible set for rs10888690 covers multiple genes, *FAF1* and *DMRTA2*; *FAF1* is involved in programmed cell death in a number of organ systems.^9^ The rs66499095 lead variant is an indel located in *TARS2*, with the credible set spanning several other genes. This variant is in high LD with several expression quantitative trait loci (eQTLs) to *TARS2*, *MRPS21*, and *RPRD2* (with one *RPRD2* eQTL in breast epithelium), and is in high LD with a plasma protein quantitative trait locus (pQTL) to *ECM1* within the African American ancestry group (LD r^2^=0.81). *TARS2* encodes a protein member of the aminoacyl-tRNA synthetase family and is linked to combination oxidative phosphorylation deficiency^10^. The rs2032905 signal is near the *TSHZ3* gene, and additionally is in high LD with eQTLs of this gene, primarily in induced pluripotent stem cells (iPSCs); this gene plays a crucial postnatal role in the function and development of corticostriatal circuitry^11^. The rs12334416 variant is located in *PINX1*, with the credible set spanning several additional genes, and is also in high LD with eQTLs to *SOX7*, *GATA4*, and *RP1L1*. *PINX1* is associated with 8p23.1 duplication syndrome linked to developmental delay^12^. The remaining signals lack coding or QTL evidence, with variant-to-gene mapping defined solely through proximity evidence: the rs35847492 credible set overlaps *TNKS*, a gene associated with 8p23.1 duplication syndrome linked to developmental delay^12^; the credible set for rs2920991 overlaps *PRAG1*, a gene that encodes an enzyme in the tyrosine kinase family^13^; the rs10852372 signal is near *C16orf72*, which has been recently identified as a regulator of telomere integrity and p53 regulation^14^; finally, the rs1855263 (∼850Kb from *DIAPH3*) and rs13421417 (∼387Kb from *LRP1B*) signals are somewhat distant to their nearest genes.

**Figure 1.**
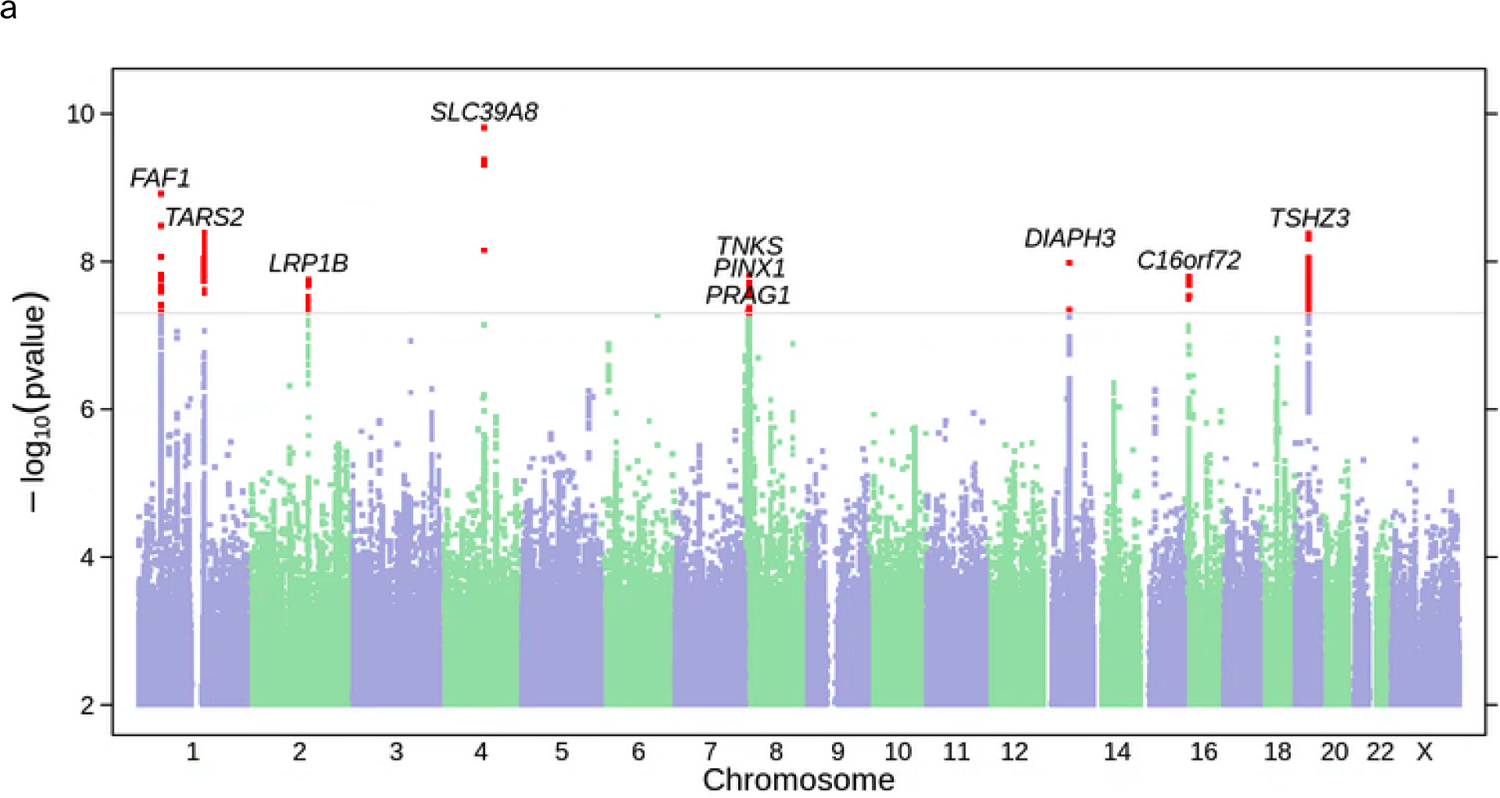
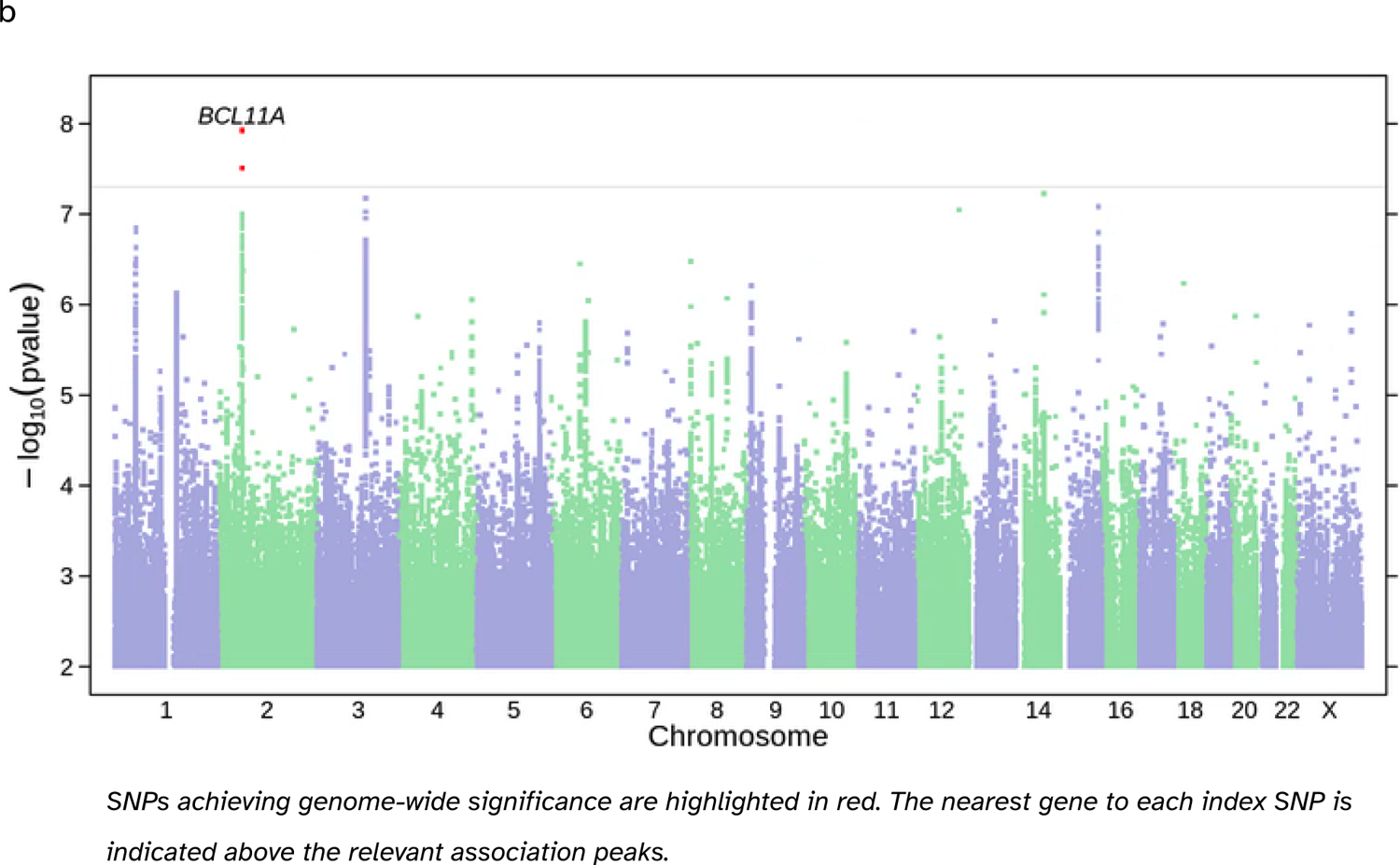
Manhattan plots showing trans-ancestral GWAS of (a) sporadic and (b) recurrent miscarriage.

**Table 2.** Loci reaching genome-wide significance for sporadic miscarriage in trans-ancestral GWAS.

| Nearby Gene | Position (GRCh38) | rsID | Effect Allele / Alternate Allele | Pooled EAF across populations | OR (95%CI) | P-value | P-value het |
| --- | --- | --- | --- | --- | --- | --- | --- |
| *SLC39A8* | Chr4:102267552 | rs13107325 | T / C | 0.758 | 1.040 (1.028 - 1.053) | 1.6e-10 | 0.704 |
| *FAF1* | Chr1:50494849 | rs10888690 | T / C | 0.570 | 0.979 (0.973 - 0.986) | 1.2e-09 | 0.708 |
| *TARS2* | Chr1:150491055 | rs66499095 | I / D | 0.804 | 0.977 (0.970 - 0.985) | 4.1e-09 | 0.282 |
| *TSHZ3* | Chr19:31414662 | rs2032905 | G / A | 0.577 | 1.020 (1.013 - 1.026) | 4.3e-09 | 0.237 |
| *DIAPH3* | Chr13:58814438 | rs1855263 | G / A | 0.672 | 1.020 (1.013 - 1.027) | 1.0e-08 | 0.796 |
| *TNKS* | Chr8:9418515 | rs35847492 | G / C | 0.707 | 0.979 (0.972 - 0.986) | 1.5e-08 | 0.318 |
| *C16orf72* | Chr16:9212279 | rs10852372 | C / A | 0.592 | 1.019 (1.012 - 1.026) | 1.6e-08 | 0.842 |
| *LRP1B* | Chr2:139844192 | rs13421417 | T / A | 0.689 | 0.981 (0.974 - 0.987) | 1.8e-08 | 0.246 |
| *PINX1* | Chr8:10739038 | rs12334416 | C / A | 0.532 | 0.982 (0.976 - 0.988) | 2.1e-08 | 0.981 |
| *PRAG1* | Chr8:8401607 | rs2920991 | T / A | 0.499 | 0.982 (0.976 - 0.989) | 4.8e-08 | 0.509 |

*P-value het derived from Cochran's Q statistical test comparing across ancestry groups.*

Of the 10 significant SNP associations identified in the trans-ancestral GWAS, six reached genome-wide significance among individuals of European ancestry whereas none reached genome-wide significance in the Latinx or African American ancestral groups (Supplementary Figure 2) with no evidence of heterogeneity of effect sizes between ancestral groups (Supplementary Table 2b).

For recurrent miscarriage, trans-ancestral GWAS of 52,087 cases and 542,880 controls identified one significant locus on chromosome 2 (rs11125830, P = 1.2e-08, OR = 1.042, 95% CI = 1.027 - 1.057, Figure 1). This variant is near a miRNA (MIR4432), and the closest protein-coding gene is *BCL11A* (∼250Kb distance), with no strong variant-to-gene mapping links. *BCL11A* functions as a transcriptional repressor that plays an important role in brain development^15^. ^16^ There was no evidence of effect size heterogeneity between individuals of differing ancestry (Cochran’s Q test p-value = 0.910; Supplementary Table 2a). Within the European ancestry GWAS, there were two additional loci (Supplementary Figure 2) that surpassed the GWAS significance threshold: chromosome 3q13.32 (rs9875219, P = 2.6e-08, OR = 1.043, 95% CI = 1.028 - 1.059), which has been previously identified within a bone development GWAS^17^, and *TENM3* (rs756996932, P = 3.1e-08, OR = 0.40, 95% CI = 0.296 - 0.536), which has been connected to syndromic microphthalmia characterized by developmental delay^18^.

In our study, recurrent miscarriage cases were nested within sporadic miscarriage cases. The variant identified from the recurrent miscarriage GWAS, rs11125830, had suggestive evidence of an association with sporadic miscarriage (P = 6.3e-05, β_sporadic miscarriage_ = 0.014, β_recurrent miscarriage_ = 0.041). The loci identified in the sporadic miscarriage analysis had the same direction of association and most had greater effect sizes for recurrent miscarriage. However, the associations of these loci with recurrent miscarriage did not reach genome-wide significance, likely due to the reduced sample size (Supplementary Table 3, Supplementary Figure 9).

The sensitivity analysis that modeled the age covariate as a spline function, made little material impact to our findings (Supplementary Figure 5, Supplementary Note). Within the analysis that restricted cases and controls to having previously reported a pregnancy, the results broadly remained consistent. The SNP rs13421417, no longer passed the threshold of GWAS significance, but rs150940764, which was identified to be in high LD (r^2^ =0.977) with rs13421417, appeared as a top hit (Supplementary Figure 4, Supplementary Figure 5). The sensitivity analysis that used age at first pregnancy as a covariate in place of age at analysis, made a negligible impact on the study. Odds ratios from the statistically significant SNPs retained consistent direction and magnitude, but had larger p-values owing to decreased sample size in the sensitivity analysis (N_cases_= 189,445, N_controls_= 260,169) compared to the main analysis (N_cases_= 284,795, N_controls_= 430,491) (Supplementary Table 10).

We sought replication using the Bonferroni-corrected level of statistical significance (P < 0.005, 0.05 / 10 tests) in the most recent European GWAS of pregnancy loss from Steinthorsdottir *et al*.^19^. Using the top hits identified within our trans-ethnic sporadic miscarriage GWAS, three variants replicated within Steinthorsdottir *et al.*’s pregnancy loss GWAS (Supplementary Table 4). The rs2920991 variant replicated at the Bonferroni-corrected threshold, while rs10852372 and rs10888690 replicated at a nominal level of significance. All tested associations were directionally consistent and had similar magnitudes of effect across the two studies.

### Observational analysis of lifestyle traits and risk of miscarriage

Previous studies have established smoking and caffeine consumption to be risk factors for miscarriage ^20–22^. Within our dataset (Supplementary Table 1), ever tobacco smokers had increased relative odds of having experienced sporadic (OR 1.16, 95%CI: 1.15 - 1.17) or recurrent (OR 1.29, 95% CI: 1.26 - 1.31) miscarriage compared to never tobacco smokers. Among smokers, each unit increase in pack years was associated with a 6% increase in relative odds of previously experiencing a sporadic miscarriage (OR 1.06; 95% CI: 1.05 - 1.07) and a 14% increase in relative odds of previous recurrent miscarriage (OR 1.14; 95%CI: 1.12 - 1.16). Being a heavy caffeine drinker was associated with a 2% increase in relative odds of previously experiencing a sporadic miscarriage (OR 1.02; 95%CI: 1.00 - 1.03) and a 4% increase in relative odds of previous recurrent miscarriage (OR 1.04; 95% CI: 1.01 - 1.07). Similarly, current caffeine consumption was associated with a higher risk of previous sporadic (OR per log10(x+75) units: 1.05 95%CI: 1.03 - 1.07) and recurrent (OR per log10(x+75) units: 1.13 95%CI: 1.09 - 1.17) miscarriage.

### Mendelian randomization (MR)

We found compelling genetic evidence in support of a causal role of liability to smoking with risk of sporadic and recurrent miscarriage. Smoking status (ever-versus never smoker) was investigated using a genetic instrument comprising 883 SNPs (mean F-statistic per SNP = 62). In the IVW MR analysis, genetic liability to ever-smoking increased the risk of both sporadic (OR 1.13; 95%CI: 1.12-1.15; P=2.61e-42) and recurrent (OR 1.25; 95%CI: 1.21-1.30; P=5.466e-34) miscarriage. Results were consistent when using robust-MR approaches with no evidence of directional horizontal pleiotropy (Figure 2, Supplementary Table 5, Supplementary Figure 6).

**Figure 2.**
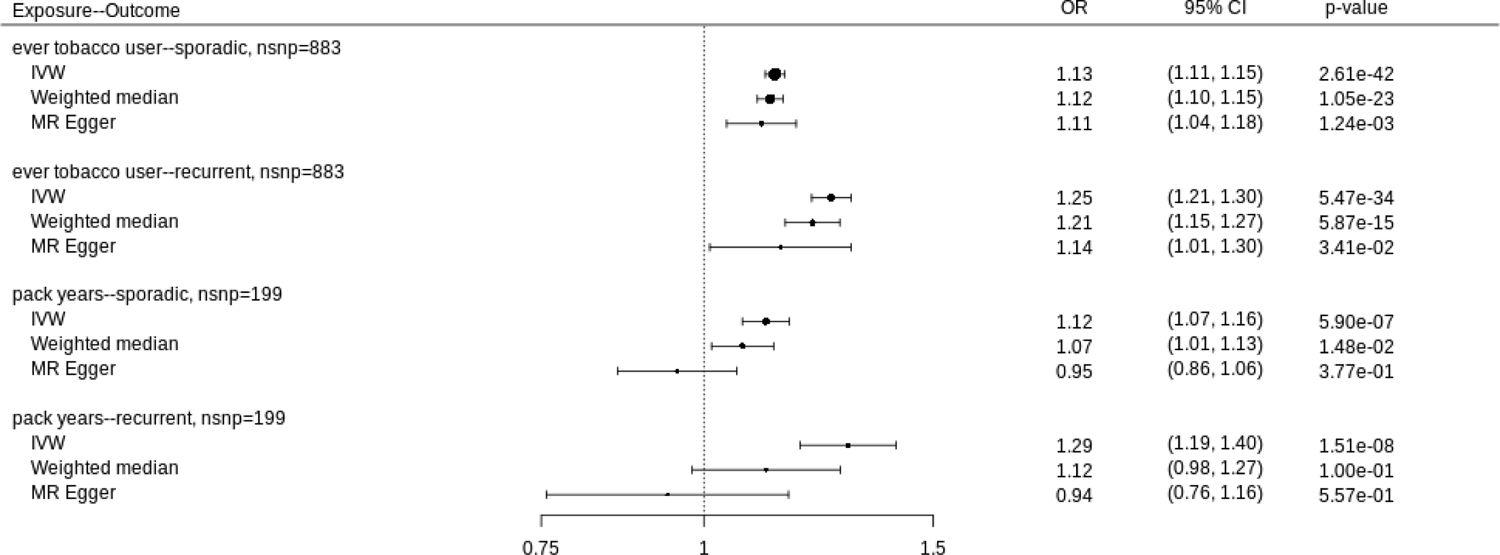
Mendelian randomization estimates of smoking and risk of sporadic and recurrent miscarriage.

Using 199 SNPs as a genetic instrument for pack years (mean F-statistic = 54) among smokers, IVW MR analysis identified a potential causal effect of higher smoking heaviness with increased risk of both sporadic (OR 1.12; 95%CI: 1.07 - 1.16; P=5.90e-7) and recurrent (OR 1.29; 95%CI: 1.19 - 1.41; P=1.514e-8) miscarriage. MR estimates trended towards the null when deploying robust MR approaches, with evidence of directional horizontal pleiotropy from MR-Egger (Figure 2, Supplementary Table 5, Supplementary Figure 6).

A potential causal role of caffeine was found. Using 127 SNPs to genetically instrument current caffeine consumption (measured in log_10_(x+75) units; mean SNP F-statistic = 73), IVW MR analysis yielded an elevated relative odds of sporadic (OR 1.27; 95%CI: 1.06 - 1.52; P=1.26e-2) and recurrent (OR 1.48; 95%CI: 1.07 - 2.04; P=1.93e-2) miscarriage, however the 95%CI were wide. Consistent effects, albeit with even wider 95% confidence intervals, were identified using robust MR approaches (Figure 3, Supplementary Table 5, Supplementary Figure 6). 113 SNPS were used as a genetic instrument for being a heavy caffeine drinker (mean F-statistic = 76). The IVW analysis identified a potential causal effect of genetic liability to heavy vs not-heavy caffeine consumption (i.e. among caffeine consumers) on the risk of sporadic (OR 1.05; 95%CI: 1.01 - 1.08; P=7.26e-3) and a directionally consistent, but more imprecise effect with risk of recurrent miscarriage (OR 1.05; 95%CI: 0.99 - 1.12 P=1.01e-1). Consistent effects, typically with wider 95%CI, were found when using robust-MR approaches (Figure 3, Supplementary Table 5, Supplementary Figure 6).

**Figure 3.**
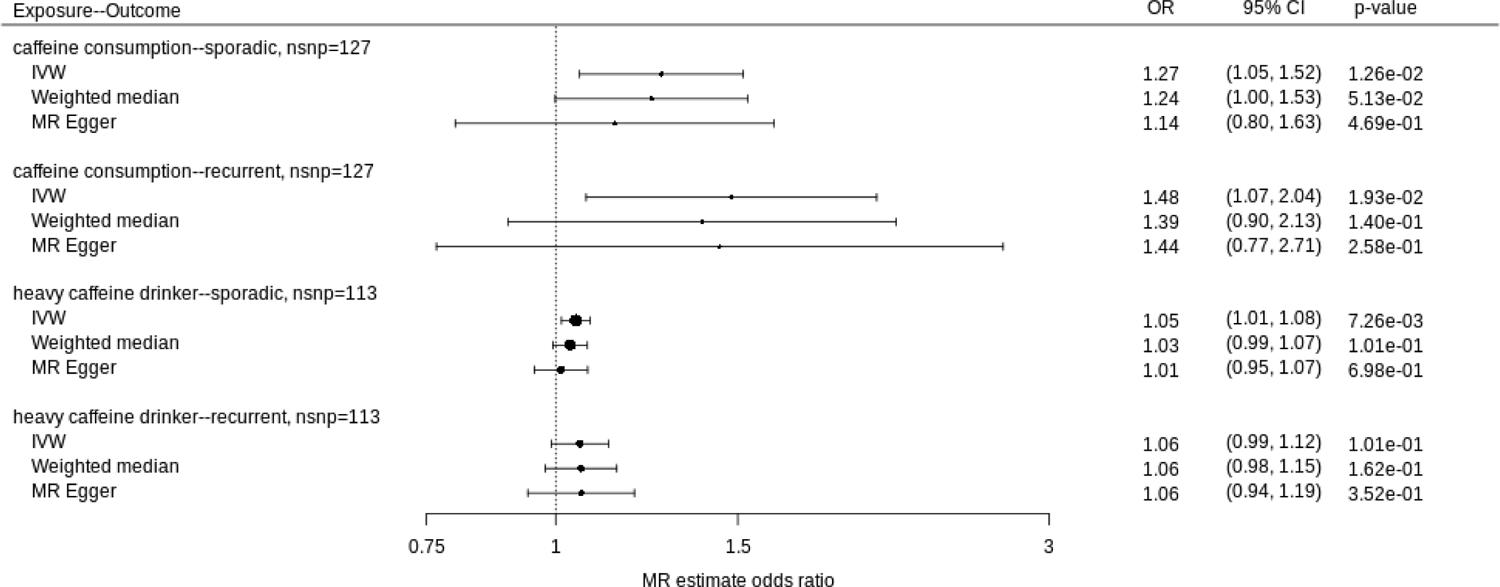
Mendelian randomization estimates of caffeine consumption and risk of sporadic and recurrent miscarriage.

Overlap in data between exposure and outcome variables is depicted in Supplementary Table 9, which showed the majority of data contributing to miscarriage overlapped with data contributing to ever tobacco smoker, with the overlap being less considerable, but still non-minimal, for the other exposure-outcome pairs. Using a split-sample design, results were broadly consistent (Supplementary Figure 7, Supplementary Figure 8, Supplementary Table 11). As a sensitivity analysis to explore potential mis-specification of the primary exposure, MR analysis using Steiger filtering yielded broadly similar findings on IVW MR (Supplementary Table 6).

## Discussion

In the largest and most diverse trans-ancestral GWAS to date, we identified 10 novel genetic loci associated with sporadic miscarriage and one genetic locus associated with recurrent miscarriage. Associated genes have roles in neural cell development, telomerase activity implicating telomere length, and conditions related to developmental delay. Mendelian randomization provided robust evidence in support of smoking and moderate evidence in support of caffeine being modifiable lifestyle causes of sporadic miscarriage.

Our study adds to the growing body of evidence of a polygenic predisposition to sporadic and recurrent miscarriage. The results of our study highlight the importance of using ancestrally diverse datasets in conducting hypothesis-free GWAS studies. This is exemplified in only a subset of the 10 loci in the trans-ancestral GWAS reaching GWAS significance among individual ancestral groups: among those of European ancestry, only 6 of the 10 SNPs reached genome-wide significance, while none did in the non-European ancestries, likely due to insufficient statistical power.

The GWAS hits from the trans-ancestral analysis are linked to biological processes that likely account for their associations with miscarriage. *TARS2* and *SLC39A8* are associated with developmental delay: *TARS2* by combined oxidative phosphorylation defect type 21, characterized by clinical manifestation within the first few months of life, resulting in severe hypotonia and early death^23^ and *SLC39A8* by hepatic manganese metabolism resulting in syndromic manifestations,^24^ mild developmental delay, and growth deficiency^25^. *TSHZ3* is associated with processes predominantly affecting neural development and plays an important role in early cortical development ^26^, with prior GWAS implicating *TSHZ3* in autism spectrum disorder ^27^. Studies in mice find TSHZ3 essential for the neuronal subpopulation in the hindbrain involved in breathing ^28^. Finally, *PINX1* enables both telomerase RNA binding activity and telomerase inhibitor activity with aberrant *PINX1* expression causing telomere shortening ^29,30^. Telomere shortening is associated with higher risk of miscarriage, as identified from a recent GWAS on pregnancy loss ^31^.

We identified one locus that reached trans-ancestral genome-wide significance with recurrent miscarriage. *BCL11A,* which functions as a transcriptional repressor that is responsible for the development of the brain, hematopoietic system development, and fetal (HbF) to adult (HbA) hemoglobin switching ^15,32,33^. *BCL11A* is highly expressed during the development of the brain and regulates developmental processes such as neuron subtype identity and, in mice, haploinsufficiency of *BCL11A* can lead to reduced brain size ^34^. Studies have shown *BCL11A* to be involved with 2p15-p16.1 microdeletion syndrome ^15^ which is characterized by developmental delay, autism spectrum disorder, microcephaly, intellectual disability, and multiple congenital organ defects ^35^. More recently, studies have identified heterozygous variants within *BCL11A* to underlie an intellectual developmental disorder, BCL11A-IDD, or Dias-Logan syndrome ^34^.

BCL11A-IDD is characterized by postnatal microcephaly, autism spectrum disorder, hypotonia, signs of autonomic dysregulation, and elevated HbF ^34^. Within the European GWAS, there were two additional SNPs that reached genome-wide significance: a SNP at chromosome 3q13.32 and *TENM3*. A *TENM3* mutation has been consistently linked to isolated or syndromic microphthalmia, anophthalmia, and coloboma ^18,36,37^.

The MR analysis robustly linked ever being a tobacco smoker to miscarriage, with analyses of smoking pack years and current caffeine consumption also suggesting potential causal roles. In the case of smoking, genetic liability to being an ever smoker elevated the risk of sporadic miscarriage by 13% and increased the risk of recurrent miscarriage by 25%. Amongst smokers, each additional pack year (equivalent to 20 cigarettes/day for a year) increased the risk of sporadic miscarriage by 12% and recurrent miscarriage by 29%, however there was evidence of directional pleiotropy for pack years, and robust MR estimates for this exposure trended towards the null, hampering robust causal deductions. Nonetheless, our findings provide reliable evidence indicative of a very probable causal effect behind tobacco smoking and miscarriage. The MR findings for tobacco are concordant with observational studies demonstrating the association between tobacco smoking and miscarriage risk. In a systematic review of 50 studies ^20^, any lifetime smoking was associated with an increased risk of miscarriage (summary relative risk = 1.23, 95% CI = 1.16 - 1.30) with the risk being greater when smoking exposure was specifically defined during pregnancy (summary relative risk = 1.32, 95% CI = 1.21 - 1.44; N = 25 studies)^20^. Our findings are also similar to other MR studies that examined the relationship between smoking and miscarriage. In a study of 259,142 individuals and 63,877 cases of pregnancy loss (defined as spontaneous abortion, stillbirth, spontaneous miscarriage, or termination), genetic predisposition to smoking initiation resulted in a combined OR for pregnancy loss of 1.31 (95% CI = 1.25–1.37)^38^. Similarly, Wang *et al*. and Laisk *et al*. both found a harmful effect between smoking and spontaneous abortion ^7,39^. The consistency of effect sizes while utilizing a larger GWAS sample size, adds to the growing literature of robust causal evidence between tobacco smoke and risk of miscarriage. Thus the totality of evidence is consistent with smoking being an important preventable cause of miscarriage.

Our findings of a possible causal role of caffeine consumption and miscarriage is of potential public health relevance, however the evidence base was less robust as for smoking, arising from less precise MR estimates. Those with genetically-predicted higher caffeine consumption had a 27% (95%CI: 5-52%) higher relative odds of sporadic and 48% (95%CI: 7-104%) higher relative odds of recurrent miscarriage. Estimates for heavy caffeine drinkers (amongst caffeine drinkers) were weaker at 5% (95%CI: 1-8%) and 6% (95%CI: −1-12%) for sporadic and recurrent miscarriage, respectively. Triangulating MR findings for caffeine consumption with contemporaneous scientific literature can help contextualize our findings. Epidemiological studies have explored the relationship between caffeine consumption (both prior to and during pregnancy) and miscarriage risk. A meta-analysis of 26 studies found caffeine consumption during pregnancy was associated with increased risk of pregnancy loss (OR 1.32, 95% CI: 1.24-1.40)^40^. Another meta-analysis found that for every 100 mg/day increment in caffeine consumption during early pregnancy, risk of miscarriage increased by 14% (95%: CI 10–19%)^41^. In a prospective cohort of pre-pregnancy individuals, those consuming >400 mg/day caffeine had a relative risk of miscarriage of 1.11 (95 % CI 0.98, 1.25) compared to individuals that consumed little caffeine each day^42^.

Dose-response relationships between the amount of caffeine consumed and miscarriage risk have also been studied. In a population based case-control study, amongst non-smokers, more miscarriages occurred in people assigned female at birth who ingested at least 100 mg of caffeine per day compared to those that drank less than 100mg, with a monotonic relationship between the amount of caffeine ingested and risk of miscarriage^43^. Therefore, a large swath of observational evidence is congruent with our MR findings, indicating that caffeine consumption may play a causal role in miscarriage. However, prior MR studies of caffeine have largely found null results ^44,38^. In the papers from Nunes *et al*.^44^, and Yuan *et al*.^38^, researchers concluded null results from the MR analysis of caffeine use and miscarriage, with directionally opposite point estimates of the odds ratios we found. The incongruence between our MR findings with those that have been previously published could be explained by our study being better statistically powered and our use of a stronger instrumental variable with 127 SNPs for caffeine consumption compared to 8 SNPs and 12 SNPs used in previous MR studies ^44,38^. Therefore, our findings are consistent with a potential causal effect of caffeine consumption on miscarriage risk. While further studies are needed to provide additional reliable evidence, the precautionary principle might suggest that individuals wishing to minimize risk of miscarriage could consider reducing their caffeine consumption.

Our study has several limitations. First, case status was ascertained through self-report, potentially introducing a source of measurement error. Second, we only evaluated the genetic contribution from genetic females, even though it is possible for genetic males to contribute towards the risk of miscarriage. Third, we were only able to partially replicate our findings in prior GWAS papers ^3,7^ perhaps due to variability in recurrent miscarriage phenotype definitions ^45^. Fourth, we did not have data on age of miscarriage however this is unlikely to impact our findings since most individuals within our cohort are past their reproductive age and were likely to have had similar ‘at-risk’ time periods for miscarriage. Fifth, in regards to the Mendelian randomization analysis, the overlapping-sample design presents a limitation. However, sensitivity analyses using a split-sample design provided congruent causal estimates ^46^. One major assumption is that the genetics of lifestyle traits at times in the life where an individual is not pregnant have relevance to when an individual is pregnant. Canalization of genetically-predicted traits such as tobacco or caffeine during pregnancy might present a unique form of bias, however the presence of such would be expected to bias MR estimates towards the null (given the “true” SNP to exposure estimate would be closer to the null in the context of such canalization). In contrast, our results are consistent with prior observational (tobacco and caffeine) and MR studies (tobacco), providing an opportunity for triangulation. Finally, in interpreting the findings of this study, we acknowledge certain limitations related to the temporal context of the data. First, similarly to other published miscarriage GWASes^7,19^, we used age at analysis rather than age at the time of miscarriage because the latter was not available. While this approach allows for some control over age-related heterogeneity, it may not fully capture the specific relationship between age and miscarriage risk. Additionally, the survey data does not establish the chronology of lifestyle factors relative to the period of miscarriage events. As such, the reported behaviors, like caffeine consumption, may not coincide with the respondents’ reproductive timelines.

In conclusion, our findings underscore a polygenic basis to sporadic and recurrent miscarriage, identifying 10 loci reaching trans-ancestral GWAS significance for sporadic miscarriage and one for recurrent miscarriage. The findings of our GWAS lay the necessary groundwork for future studies of miscarriage among non-Europeans, given the higher risk of miscarriage among these populations ^2,47^. We find strong evidence that smoking is etiologically detrimental to miscarriage and that caffeine may also be of causal relevance. Future studies might explore risk stratification and genetic epidemiological approaches to further characterize etiology to help individuals identify their potential risk of experiencing a miscarriage, ultimately, reducing morbidity around this common adverse outcome of pregnancy.

## Data Availability

The full GWAS summary statistics for the 23andMe discovery data set will be made available through 23andMe to qualified researchers under an agreement with 23andMe that protects the privacy of the 23andMe participants. Datasets will be made available at no cost for academic use. Please visit https://research.23andme.com/collaborate/#dataset-access/ for more information and to apply to access the data.

## Acknowledgements

We would like to thank the research participants and employees of 23andMe for making this work possible. The following members of the 23andMe Research Team contributed to this study: Adam Auton, Elizabeth Babalola, Robert K. Bell, Ninad S. Chaudhary, Zayn Cochinwala, Sayantan Das, Emily DelloRusso, Sarah L. Elson, Nicholas Eriksson, Chris Eijsbouts, Teresa Filshtein, Pierre Fontanillas, Davide Foletti, Will Freyman, Zach Fuller, Julie M. Granka, Chris German, Éadaoin Harney, Alejandro Hernandez, Barry Hicks, David A. Hinds, Reza Jabal, Ethan M. Jewett, Yunxuan Jiang, Matt Kmiecik, Katelyn Kukar, Alan Kwong, Keng-Han Lin, Yanyu Liang, Bianca A. Llamas, Aly Khan, Steven J. Micheletti, Matthew H. McIntyre, Meghan E. Moreno, Dominique T. Nguyen, Jared O’Connell, Steve Pitts, G. David Poznik, Morgan Schumacher, Leah Selcer, Anjali J. Shastri, Shubham Saini, Suyash Shringarpure, Qiaojuan Jane Su, Joyce Y. Tung, Susana A. Tat, Vinh Tran, Xin Wang, Wei Wang, Catherine H. Weldon, Peter Wilton. A.R., P.N., J.S., M.V.H., and S.A. are employed by and hold stock or stock options in 23andMe, Inc.

## Methods

### Study participants and data collection

This study utilizes self-reported data collected through a web-based fertility survey fielded on 23andMe’s web platform. As part of the 23andMe service, customers are genotyped on SNP microarrays and offered the opportunity to participate in scientific research; approximately 80% of customers consent to do so. Individuals participating in this study provided informed consent and conducted survey participation online, under a protocol approved by the external AAHRPP-accredited IRB, Ethical & Independent Review Services (E&I Review, now Salus IRB).

23andMe research participation is generally conducted via online surveys. The fertility survey was open to all consented research participants aged 18 or older. Consented participants who previously reported a diagnosis or diagnoses of infertility or subfertility, preeclampsia, endometriosis, uterine fibroids, polycystic ovarian syndrome, gestational diabetes, or cholestasis were more likely to be surfaced the survey compared to those who had not previously reported a diagnosis or diagnoses of the aforementioned conditions. This resulted in a survey sample enriched for infertility conditions.

Within the fertility survey, participants were asked about their reproductive health history, including fertility treatments, time to conception, miscarriage occurrence, full term births, fertility related surgeries, and hormone levels. Additional participant information, including age, lifestyle characteristics, and preexisting health conditions, were gathered through previous 23andMe surveys. Analysis was restricted to genetic females (XX) between the ages of 18 and 85 inclusive, had non-missing data on self-reported miscarriage, and had non-missing data for the covariates within the modeling analyses.

### Case definitions

For the purposes of this study, we defined sporadic miscarriage in accordance with the World Health Organization (WHO) definition on the basis of a spontaneous loss of an embryo or fetus ^48^. Recurrent miscarriage, seen as a more severe case definition, was defined within this study as the loss of three or more pregnancies ^48^.

Individuals with existing data on miscarriage were categorized into three groups based on how they responded to questions related to miscarriage. Participants who answered “yes” to “Have you ever experienced a miscarriage?” or answered a numerical value of 1 or more to the question “How many miscarriages have you ever had?” were considered a sporadic miscarriage case. Participants who answered “yes” and a value greater than or equal to 3, were considered a recurrent miscarriage case. Thus recurrent miscarriage cases were nested among individuals who were ascertained as having a sporadic miscarriage. If an individual replied “no” or a value of 0 to the above questions, they were assigned as a control (Supplementary Figure 1).

### Descriptive analysis

Demographic, behavioral, and health characteristics were collected from participants (Table 1). Age was recorded as each research participant’s age at the time of analysis. Ancestry was inferred via a previously described genetic ancestry classification algorithm^49^. Tobacco smoking was dichotomized by a smoking history of at least 100 cigarettes in a lifetime or reported being a regular user (yes/no). Our tobacco smoking variable followed the established standard definition used by the US Centers for Disease Control and Prevention (CDC) in their National Health Interview Survey ^50^. Caffeine consumption was measured as daily milligrams of caffeine from any of the following: coffee, tea, soda, or energy drinks. Caffeine consumption was then transformed by log_10_(x+75) to create a more normally distributed variable. We collected data on the following reproductive organ diseases: polycystic ovary syndrome (PCOS), preeclampsia, hyperandrogenism, endometriosis, ovarian cysts, and uterine fibroids. Data collection surrounding these variables followed a similar format where participants were asked if they have been diagnosed with, or treated for, the aforementioned conditions (yes/no). All variable definitions asked within the survey are reported in the Supplementary Note.

### Genotyping and SNP Imputation

DNA extraction and genotyping were performed on saliva samples by Clinical Laboratory Improvement Amendments-certified and College of American Pathologists-accredited clinical laboratories of Laboratory Corporation of America. Participants were genotyped on one of five genotyping platforms. The v1 and v2 platforms were based on the Illumina HumanHap550+ BeadChip, including about 25,000 custom variants selected by 23andMe, with a total of about 560,000 SNPs. The v3 platform was based on the Illumina OmniExpress+ BeadChip, with custom content to improve the overlap with our v2 array, with a total of about 950,000 variants. The v4 platform was a fully customized array, including a lower redundancy subset of v2 and v3 variants with additional coverage of lower-frequency coding variation, and about 570,000 variants. The v5 platform, in current use, is an Illumina Infinium Global Screening Array (∼640,000 variants) supplemented with ∼50,000 variants of custom content. This array was specifically designed to better capture global worldwide genetic diversity and to help standardize the platform for genetic research. Samples that failed to reach 98.5% call rate were re-genotyped. A total of 1,522,458 variants have been genotyped across the five genotyping platforms and 1,469,237 variants produced analyzable results.

Variants were imputed using three combined independent reference panels: the publicly available Human Reference Consortium (HRC), and UK BioBank (UKBB) 200K Whole Exome Sequencing (WES) reference panels and the 23andMe reference panel, which was built by 23andMe using internal and external cohorts. The publicly available HRC data were downloaded from the European Genome-Phenome Archive and includes 27,165 samples. Variants were liftovered to hg38 and excluded if their new positions were on a different chromosome. Variants were then re-phased using SHAPEIT4 ^51^. Singletons were then excluded. The final HRC reference panel included 27,165 samples and 39,057,040 SNPs (no indels). The publicly available UKBB 200K WES data includes 200,643 whole exome sequenced samples (mostly British ancestry) and more than 22M variants ^52^. After applying the following filters: multi-allelics were split into bi-allelic variants (with bcftools), genotypes with GQ<20 were set to missing, variants with >20% missingness were removed, variants with minor allele count == 0 were removed, variants with inbreeding coefficient < −0.3 (high heterozygosity) were removed, 17,975,023 variants were available. After excluding singletons and applying relevant QCs, the UKBB 200K WES reference panel included 199,815 samples and 17,082,588 variants (16,115,376 SNPs and 967,212 indels). The final 23andMe reference panel included 12,217 samples and 82,078,539 variants (73,852,355 SNPs + 8,226,184 indels). The final imputation panel included a total of 99,675,338 variants (90,582,19 SNPs and 9,093,144 indels). Because participants were genotyped on one of the five genotyping platforms (v1 to v5), the imputation is performed independently for each genotyped platform.

### Ancestry Classification

A detailed description of the 23andMe ancestry classifier can be found here ^53^. Briefly, 23andMe ancestry classifier algorithm determines participant ancestries through an analysis of local ancestry ^54^. It first partitions phased genotyped data into short windows of about 300 SNPs. Within each window, we use a support vector machine (SVM) to classify individual haplotypes into one of 45 worldwide reference populations. The SVM classifications are then fed into a hidden Markov model (HMM) that accounts for switch errors and incorrect assignments, and gives probabilities for each reference population in each window. Finally, we used simulated admixed individuals to recalibrate the HMM probabilities so that the reported assignments are consistent with the simulated admixture proportions.

### GWAS

A maximal set of unrelated individuals was chosen for each GWAS analysis using a segmental identity-by-descent (IBD) estimation algorithm ^55^. Participants were defined as related if they shared more than 700 cM IBD, including regions where the two individuals shared either one or both genomic segments IBD. When selecting individuals for case/control phenotype analyses, the selection process was designed to maximize case sample size by preferentially retaining cases over controls. Specifically, if both an individual case and an individual control were found to be related, then the case was retained in the analysis.

We then computed association test results for the genotyped and the imputed SNPs. For case control phenotypes, we computed associations by logistic regression assuming additive allelic effects. For tests using imputed data, we used the imputed dosages rather than best-guess genotypes. As standard, we included covariates for age and the top five principal components to account for residual population structure, and indicators for genotype platforms to account for genotype batch effects. The association test P value we report is computed using a likelihood ratio test, which in our experience is better behaved than a Wald test on the regression coefficient.

GWAS summary statistics from genotyped and imputed data were then combined, favoring imputed results when choosing between imputed and genotyped results, unless the imputed variant was unavailable or failed quality control. For the imputed results, we removed variants with low imputation quality (r^2^ < 0.5 averaged across batches or a minimum r^2^ < 0.3) or with evidence of batch effects (analysis of variance (ANOVA) F-test across batches, P < 10^−50^). For the genotyped variants, we removed variants only present on our V1 or V2 arrays (due to small sample size) that failed a Mendelian transmission test in trios (P < 10^−20^), failed a Hardy–Weinberg test in individuals of European ancestry (P < 10^−20^), failed a batch effect test (ANOVA P < 10^−50^) or had a call rate <90%.

We repeated the GWAS analysis for both phenotypic traits separately within each genetic ancestral population for which we had sufficient sample size: European, Latinx, and African American (Supplementary Figure 2). The resulting summary statistics were corrected for inflation using LD score regression intercept for our European dataset, and genomic control for Latinx and African American datasets, when the inflation factor was estimated to be greater than 1(λ_Latinx sporadic miscarriage_=1.044, λ_African American sporadic miscarriage_ =1.045, λ_Latinx recurrent miscarriage_ =1.018, λ_African American recurrent miscarriage_=0.997).

A trans-ancestral meta-analysis of the three populations was then performed using a fixed effect model (inverse variance weighted method), restricting to variants of at least 20 minor allele count in each constituent population and again corrected for inflation using genomic control (inflation factor for sporadic miscarriage = 1.022, inflation factor for sporadic miscarriage = 1.084). For the trans-ancestral meta-analysis, an associated locus was defined by identifying all variants with meta-analysis P-value < 1e-5, by grouping these variants into intervals separated by gaps of at least 250 kb, and by choosing the variant with smallest P-value within each interval.

### Functional annotation of GWAS index variants

To perform variant-to-gene mapping, hypotheses of functionally relevant genes are generated by annotating the strongest associations (index variants) with nearby functional variants. The mapping is computed by searching functional variants within 500 kb of the index variant with a filter of linkage disequilibrium r2 > 0.8. Functional variants for mapping include coding variants (annotated by the Ensembl Variant Effect Predictor (VEP) v109 ^56^), eQTLs, and pQTLs. The eQTL annotation resources consist of a comprehensive collection of standardized variants impacting gene expression in various tissues obtained from publicly available datasets ^57,58,59,60,61,62,63,64,65,66,67,68,69^ and datasets processed by the 23andMe eQTL pipeline. The pQTL annotation resources similarly include a collection of curated protein QTLs from relevant public datasets ^70^.

### eQTL discovery

eQTL calling was performed with one of two versions of 23andMe pipelines, depending on the dataset in question (Supplementary Table 12). The first pipeline used FastQTL^71^ in permutation mode, restricting all tests to variants within a window defined to be 1Mbp up- or downstream of a given gene’s transcription start site (TSS). Variants tested were single nucleotide polymorphisms with an in-sample MAF ≥ 1%, to avoid errors in detection or mapping of larger genetic variants in cross-ancestry comparisons, and models were adjusted for age (if available), sex, probabilistic estimation of expression residuals (PEER) factors^72^, genetic PCs, and per-dataset covariates. For each gene, the index variant was identified by the minimal permutation p-value. eQTLs were called then on lead variants if they passed a 5% FDR filter using Storey’s q-value ^73^ methodology.

Conditional eQTLs were identified via FastQTL’s permutation mode, by using each eQTL as an additional covariate in the model for a given gene. The lead conditional eQTL for all genes were again FDR controlled at 5%, and a maximum of 10 conditional steps were run. Finally, for a set of conditional eQTLs for a given gene, a joint model was fit, and the final eQTL callset consisted of those eQTLs whose joint model test passed a 5% FDR filter. eQTLs were only called for genes classified as one of ‘protein_coding’, ‘miRNA’, ‘IG_C_gene’, ‘IG_D_gene’, ‘IG_J_gene’, ‘IG_V_gene’,’TR_C_gene’, ‘TR_D_gene’, ‘TR_J_gene’, ‘TR_V_gene’ as defined in GENCODE^74^.

The second version of the 23andMe pipeline uses strand-aware RNA-seq quantification, and the eQTLs were called using SusieR package^75^ instead of FastQTL, with expression PCs (selected with the elbow method) replacing PEER factors in the modeling, and using the GENCODE v43 gene model (Supplementary Table 12). The pipeline natively generated credible sets with a set probability to contain a SNP tagging the causal variant.

### GWAS sensitivity analyses

We conducted three sensitivity GWAS analyses. First, to ensure controls represented the population that gave rise to the cases, cases and controls were required to have reported being pregnant. Second, we modeled age as a spline function as previous studies demonstrated potential non-linear relationships between miscarriage and age ^76^. Further rationale to conduct this sensitivity analysis can be found in the Supplementary Note. Third, a regression was run on the top hits derived from the European ancestry sporadic miscarriage GWAS using age at first pregnancy as a covariate into the model rather than age at analysis.

### Mendelian randomization analysis

We deployed a Mendelian randomization (MR) analysis in European individuals to examine the potential causal relationships between tobacco smoking and caffeine consumption and risk of sporadic and recurrent miscarriage. The MR analysis was performed using an overlapping-sample design, in accordance with published methods proposed by Sanderson *et al.* to account for potential horizontal pleiotropy and potential bias due to overlapping samples ^77^. The split-sample design used the following sample generating schema: individuals were filtered for relatedness with a preference towards keeping those with miscarriage data over those with exposure data, given that most individuals with miscarriage data also had exposure data. All individuals with miscarriage data were then included into one set, and all individuals without miscarriage data were put into the exposure set. To obtain a more comprehensive understanding behind miscarriage risk and lifestyle factors, multiple caffeine and smoking variables were examined along with the ever tobacco smoker and current caffeine variables described in the Methods section above. The other variables of interest included: tobacco smoking pack years and a binarized heavy caffeine drinker variable. Participants self-reported a quantitative measure of the amount of tobacco cigarettes smoked, using a calculated pack years. One pack year is the equivalent to smoking 20 cigarettes per day for 365 days. Participants also self-reported their current caffeine consumption and were dichotomized into being a “heavy caffeine drinker” if they previously reported drinking several caffeinated beverages per day, or “not a heavy caffeine drinker” if reported less than that amount.

Association tests were run on exposures of interest. The exposure variables of interest were modeled as follows: pack years and current caffeine consumption were modeled as continuous variables, while ever being tobacco smoker and being a heavy caffeine drinker were modeled as binary variables. All variables were recorded cross-sectionally, which for most participants was after they had experienced a sporadic or recurrent miscarriage. As such, the analysis is limited to exploring associations in cross-sectional data rather than examining temporal associations. In each model, age (determined at the time of analysis), and ancestry (self-reported ethnicity determined at the time of analysis) were included as covariates. Models all followed a similar form: miscarriage ∼ age + ancestry + potential risk factor (ever tobacco smoker, heavy caffeine drinker, pack years, or current caffeine consumption). Odds ratios (OR) and 95% confidence intervals (CI) were calculated for each of the variables in the models. Since we ran a total of 4 tests we used a Bonferroni adjusted p-value of 0.0125 (0.05/4 tests) as the threshold for statistical significance.

To obtain MR estimates of lifestyle behaviors for sporadic and recurrent miscarriage, several MR approaches were employed. The analysis, via the TwoSampleMR R package^78^, ran three different methods. The random effects IVW analysis, which assumes no horizontal pleiotropy, used a linear regression constrained to the origin of variant-exposure and the variant-outcome associations for each instrument, weighted by their inverse variance. Two robust MR methods were also used: the weighted median method which assumes that more than 50% of the weight in the analysis originates from valid instruments, and MR-Egger analysis was used to explore, and potentially correct for the presence of horizontal pleiotropy, and the results of the analysis were presented as odds ratios and 95% confidence intervals which provides an estimate of relative risk. MR scatter plots presented in Supplementary Figure 6, were generated using mr() and mr_scatter_plot() from the TwoSampleMR R package.

### Instrument construction for MR

We opted to use 23andMe exposure data to synthesize genetic instruments for each trait of interest rather than external data because of the large available sample sizes and resulting statistical power. Summary statistics of each lifestyle variable were obtained from the most recent GWAS for these traits, run in-house. SNPs associated with each of the risk factors (tobacco smoker, smoking in pack years, heavy caffeine drinker, and current caffeine consumption) were included as instrumental variables if they met a genome-wide significance level of p< 5e–8. The strength of the instruments were assessed using the mean F statistic, as calculated by the system_metrics function in the TwoSampleMR R package^78^.

### Statistical analysis

All statistical analyses were performed in Rstudio version 4.1.2 and were run using rlib23, a proprietary 23andMe software package.

