## Supplementary_Material for "Trans-ancestral Genome Wide Association Study of Sporadic and Recurrent Miscarriage"

### Supplementary Note


*Additional Variable Definitions*

*Sensitivity Analysis*

Supplementary Table 1- *Summary of Conventional Observational Association Analyses*

Supplementary Table 2- *Genetic associations and odds ratios within the recurrent and sporadic miscarriage variable*

Supplementary Table 3- *Evaluating effect sizes between recurrent and sporadic miscarriage in trans-ancestral GWAS results*

Supplementary Table 4- *Look-ups of trans-ancestral sporadic miscarriage GWAS hits in our study in a consortium paper's sporadic GWAS summary statistics (Laisk et al.)*

*Supplementary Table 5- Intercepts and 95% CIs from MR analysis*

*Supplementary Table 6- Steiger filtered Random effects IVW results from Mendelian Randomization analysis*

*Supplementary Table 7- Variant-to-gene mapping (LD rsqr > 0.8 coding, eQTL, or pQTL variants) for for sporadic miscarriage per population*

*Supplementary Table 8- EAFs for sporadic miscarriage top hits per ancestry group*

*Supplementary Table 9- Sample overlap for each exposure and outcome for Mendelian Randomization analysis*

*Supplementary Table 10- Rerun of European sporadic miscarriage GWAS Top Hits, using age at first pregnancy as a covariate*

*Supplementary Table 11- Intercepts and 95% CIs from split-sample MR analysis*

*Supplementary Table 12- Public datasets that are internally processed using 23andme eQTL pipelines*

Supplementary Figure 1- *Study flowchart depicting how miscarriage cases and controls were enrolled into study*

Supplementary Figure 2- *Manhattan plots for both recurrent and sporadic miscarriage variables and each population prior to trans-ancestral meta-analysis*

Supplementary Figure 3- *Locus zoom plots for SNPs that reached genome-wide significance on trans-ancestral analysis of sporadic miscarriage and recurrent miscarriage*

Supplementary Figure 4- *Forest plots of SNPs reaching GWAS significance for sporadic miscarriage comparing main analysis to sensitivity analysis*

Supplementary Figure 5- *Manhattan plots of sensitivity analyses*

*Supplementary Figure 6- MR Scatter plots*

*Supplementary Figure 7- Split-sample Mendelian randomization estimates of smoking and caffeine intake and risk of sporadic and recurrent miscarriage*

*Supplementary Figure 8- MR Scatter plots for split-sample design*

*Supplementary Figure 9- Effect size forest plots for trans-ancestral sporadic and recurrent miscarriage top hits*

*Additional Variable Definitions*

- Education status was recorded as the highest degree or level of school a research participant has completed.
- Body mass index (BMI) was extracted from self-reported height and weight questions then calculated from mass (kg) divided by height squared ($m^2$).
- Daily alcohol consumption was extracted from self-reported daily alcohol beverage consumption (in servings).
- Age of menarche was recorded as the age an individual first started their menstrual period.
- Number of biological children was recorded as the number of biological children an individual has, with individuals with no children being included into the variable.
- The number of pregnancies was recorded as the number of times a research participant reported being pregnant with options being: ‘once’, ‘twice’, ‘three times’, ‘four times’, and ‘more than five times’. This variable was then converted into a numeric variable with the maximum value being 5.
- Age of first pregnancy was recorded by collecting data regarding the age at the first time an individual became pregnant.

*Sensitivity Analysis*

In a separate study investigating the role of maternal age and risk of miscarriage, researchers concluded there is a strong non-linear influence of maternal age, and showed age associated risk of miscarriage follows a J-shaped curve [1]. To mimic this trend, we modeled age as a spline to add more granularity and capture the complex relationship between age and risk of miscarriage.

Restricting cases and controls to having previously reported if they experienced a pregnancy had only a minor impact on our findings (Supplementary Figure 5). For our recurrent miscarriage analysis, we still identified one locus near the gene, *BCL11A*. The

P-value of the genetic variant remained consistent (p-value $_{\text{original analysis}}$ = 1.2e-08, p-value $_{\text{sensitivity analysis}}$ = 1.5e-08) . For the sporadic miscarriage analysis, most of the genes retained genome-wide significance, with a new hit at *PKHD1L1* (p-value $_{\text{original analysis}}$ = 1.3e-07, p-value $_{\text{sensitivity analysis}}$ = 9.7e-09). Additionally, the previously significant SNP on chr16p13.2 no longer surpassed the threshold of significance (p-value $_{\text{original analysis}}$ = 1.6e-08, p-value $_{\text{sensitivity analysis}}$ = 1.5e-07). *TSHZ3* was observed to have a smaller P-value (p-value $_{\text{original analysis}}$ = 4.3e-09, p-value $_{\text{sensitivity analysis}}$ = 3.3e-10, Supplementary Figure 5). Sample size for the sensitivity analysis had a consistent reduction across both phenotypes (N $_{\text{SM original analysis}}$ = 839,601, N $_{\text{SM sensitivity analysis}}$ = 820,001, N $_{\text{RM original analysis}}$ = 594,967, N $_{\text{RM sensitivity analysis}}$ = 575,353).

Modeling the covariate age as a spline function, we observed no major changes to our findings. All GWAS hits for sporadic miscarriage remained significant, with the addition of an additional locus that met the threshold of significance near chr6q22 (p-value = 4.49e-08). For analyses using the recurrent miscarriage variable *BCL11A* remained the only gene that passed the threshold of genome-wide significance.

**Supplementary Table 1: Summary of Conventional Observational Association Analyses**

| | Sporadic Micarriage Odds ratio (95%CI) | Recurrent Miscarriage Odds ratio (95%CI) |
|---|---|---|
| **Tobacco smoker (ever smoker versus never)** | 1.16 (1.15 - 1.17)** | 1.29 (1.26 - 1.31)** |
| **Smoking pack years † (in pack years)** | 1.06 (1.05 - 1.07)** | 1.14 (1.12 - 1.16)** |
| **Current caffeine consumption † † (log10(x+75), in mg)** | 1.05 (1.03 - 1.07)** | 1.13 (1.09 - 1.17)** |
| **Heavy caffeine drinker (heavy caffeine drinker versus less caffeine drinker)** | 1.02 (1.00 - 1.03)** | 1.04 (1.01 - 1.07)** |

*Supplementary Table 1: Symbols correspond to the P significance level; *P < 0.05, **P<0.0125. Logistic regressions were minimally adjusted for age at survey and ancestry. † Pack years is amongst smokers only. † † Current caffeine consumption includes individuals that do not consume caffeine. 95%CI = 95% confidence interval*

**Supplementary Table 2a: Genetic associations and odds ratios within the recurrent miscarriage variable near BCL11A (rs11125830)**

| Effect Allele | Population | MAF, controls | MAF, recurrent miscarriage cases | OR | p-value | Cochran's Q p-value |
|---|---|---|---|---|---|---|
| G | European | 0.566 (N = 465,220) | 0.577 (N = 44,468) | 1.043 | 5.92e-09 | 0.910 |
| | Latinx | 0.628 (N = 59,363) | 0.633 (N = 5,701) | 1.047 | 0.276 | |
| | African American | 0.704 (N = 18,297) | 0.708 (N = 1,918) | 1.026 | 0.508 | |

*MAF = minor allele frequency; OR = odds ratio. Cochran's Q p-value is derived from Cochran's Q statistical test comparing across ancestry groups.*

**Supplementary Table 2b: Genetic associations and odds ratios within the sporadic miscarriage variables**

| rsID | Effect Allele | Population | MAF, controls | MAF, sporadic miscarriage cases | OR | p-value | Cochran's Q p-value |
|---|---|---|---|---|---|---|---|
| | | European | 0.080 (N = 430,491) | 0.083 (N = 284,795) | 1.039 | 5.92e-09 | |
| rs13107325 | T | Latinx | 0.061 (N = 56,436) | 0.063 (N =37,561) | 1.051 | 0.0157 | 0.704 |
| | | African American | 0.023 (N = 18,081) | 0.025 (N = 12,237) | 1.079 | 0.186 | |
| | | European | 0.586 (N = 430,491) | 0.580 (N = 284,795) | 0.979 | 2.45e-09 | |
| rs10888690 | T | Latinx | 0.633 (N = 56,436) | 0.627 (N =37,561) | 0.969 | 0.112 | 0.708 |
| | | African American | 0.462 (N = 18,081) | 0.459 (N = 12,237) | 0.990 | 0.570 | |
| | | European | 0.784 (N = 430,491) | 0.780 (N = 284,795) | 0.976 | 5.95e-09 | |
| rs66499095:CA_C | I | | | | | | 0.282 |

| | | | | | | | |
|---|---|---|---|---|---|---|---|
| | | Latinx | 0.838 (N = 56,436) | 0.834 (N =37,561) | 0.974 | 0.0490 | |
| | | African American | 0.698 (N = 18,081) | 0.698 (N = 12,237) | 1.006 | 0.759 | |
| rs2032905 | G | European | 0.601 (N = 430,491) | 0.605 (N = 284,795) | 1.018 | 5.31e-07 | 0.237 |
| | | Latinx | 0.565 (N = 56,436) | 0.573 (N =37,561) | 1.036 | 4.01e-4 | |
| | | African American | 0.382 (N = 18,081) | 0.383 (N = 12,237) | 1.012 | 0.492 | |
| rs1855263 | G | European | 0.669 (N = 430,491) | 0.673 (N = 284,795) | 1.019 | 2.94e-07 | 0.796 |
| | | Latinx | 0.671 (N = 56,436) | 0.676 (N =37,561) | 1.023 | 0.0272 | |
| | | African American | 0.604 (N = 18,081) | 0.610 (N = 12,237) | 1.030 | 0.0945 | |
| rs35847492 | G | European | 0.683 (N = 430,491) | 0.678 (N = 284,795) | 0.980 | 4.61e-08 | 0.318 |

| | | | | | | | |
|---|---|---|---|---|---|---|---|
| | | African American | 0.807 (N = 18,081) | 0.800 (N = 12,237) | 0.959 | 0.0513 | |
| rs10852372 | C | European | 0.591 (N = 430,491) | 0.596 (N = 284,795) | 1.018 | 2.97e-07 | 0.842 |
| | | Latinx | 0.563 (N = 56,436) | 0.569 (N =37,561) | 1.024 | 0.0146 | |
| | | African American | 0.844 (N = 18,081) | 0.849 (N = 12,237) | 1.022 | 0.382 | |
| rs13421417 | T | European | 0.663 (N = 430,491) | 0.658 (N = 284,795) | 0.980 | 1.50e-08 | 0.246 |
| | | Latinx | 0.731 (N = 56,436) | 0.729 (N =37,561) | 0.995 | 0.633 | |
| | | African American | 0.886 (N = 18,081) | 0.883 (N = 12,237) | 0.953 | 0.0868 | |
| rs12334416 | C | European | 0.542 (N = 430,491) | 0.538 (N = 284,795) | 0.982 | 1.82e-07 | 0.981 |
| | | Latinx | 0.500 (N = 56,436) | 0.495 (N =37,561) | 0.982 | 0.0608 | |

| | | | | | | | |
|---|---|---|---|---|---|---|---|
| | | African American | 0.653 (N = 18,081) | 0.649 (N = 12,237) | 0.979 | 0.232 | |
| rs2920991 | T | European | 0.502 (N = 430,491) | 0.497 (N = 284,795) | 0.981 | 3.87e-08 | 0.509 |
| | | Latinx | 0.488 (N = 56,436) | 0.487 (N =37,561) | 0.993 | 0.470 | |
| | | African American | 0.729 (N = 18,081) | 0.727 (N = 12,237) | 0.980 | 0.302 | |

*MAF = minor allele frequency; OR = odds ratio. Cochran's Q p-value is derived from Cochran's Q statistical test comparing across ancestry groups.*

**Supplementary Table 3: Evaluating effect sizes between recurrent and sporadic miscarriage in trans-ancestral GWAS results**

| rsID | Sporadic Miscarriage trans-ancestral GWAS | | | Recurrent Miscarriage trans-ancestral GWAS | | | Z-test p-value |
|---|---|---|---|---|---|---|---|
| | Effect Size | 95% CI | p-value | Effect Size | 95% CI | p-value | |
| *SNPs associated with recurrent miscarriage* | | | | | | | |
| rs11125830 | 0.014 | (0.0069 - 0.020) | 6.3e-05 | 0.041 | (0.027 - 0.056) | 1.2e-08 | 5.4e-4** |
| *SNPs associated with sporadic miscarriage* | | | | | | | |
| rs13107325 | 0.039 | (0.027 - 0.052) | 1.6e-10 | 0.046 | (0.021 - 0.072) | 3.3e-4 | 6.3e-1 |
| rs10888690 | -0.021 | (-0.028 - -0.014) | 1.2e-09 | -0.036 | (-0.049 - -0.021) | 7.9e-7 | 6.4e-2** |
| rs66499095 | -0.023 | (-0.031 - -0.016) | 4.1e-09 | -0.041 | (-0.057 - -0.025) | 7.5e-7 | 5.4e-2 |
| rs2032905 | 0.020 | (0.013 - 0.026) | 4.3e-09 | 0.024 | (0.011 - 0.038) | 5.3e-4 | 5.4e-1 |
| rs1855263 | 0.020 | (0.013 - 0.027) | 1.0e-08 | 0.032 | (0.017 - 0.046) | 1.4e-5 | 1.4e-1 |
| rs35847492 | -0.021 | (-0.028 - -0.014) | 1.5e-08 | -0.019 | (-0.034 - -0.0036) | 1.5e-2 | 8.1e-1 |
| rs10852372 | 0.019 | (0.012 - 0.025) | 1.6e-08 | 0.028 | (0.014 - 0.042) | 6.0e-5 | 2.3e-1 |
| rs13421417 | -0.019 | (-0.026 - -0.013) | 1.8e-08 | -0.014 | (-0.029 - -0.0001) | 4.8e-2 | 5.3e-1 |
| rs12334416 | -0.018 | (-0.025 - -0.012) | 2.1e-08 | -0.026 | (-0.040 - -0.013) | 1.3e-4 | 2.9e-1 |
| rs2920991 | -0.018 | (-0.024 - -0.011) | 4.8e-08 | -0.019 | (-0.033 - -0.0059) | 4.8e-3 | 8.4e-1 |

*** denotes if the rsID has reached nominal significance ($P < 0.05$). Z-test is used to evaluate heterogeneity between sporadic and recurrent miscarriage estimates. 95%CI = 95% confidence interval. Effect sizes refer to beta coefficients on the log scale.*

**Supplementary Table 4: Look-ups of trans-ancestral sporadic miscarriage GWAS hits in our study in a previously published pregnancy loss GWAS summary statistics (Steinthorsdottir et al.)**

<table>
<tr><th rowspan="2">rsID</th><th colspan="4">Our Study</th><th colspan="3">Steinthorsdottir et al.</th></tr>
<tr><th>Pooled EAF across populations</th><th>OR</th><th>95% CI</th><th>p-value</th><th>EA (EAF)</th><th>OR</th><th>p-value</th></tr>
<tr><td>rs13107325</td><td>T (0.758)</td><td>1.040</td><td>(1.028 - 1.053)</td><td>1.6e-10</td><td>T (0.052)</td><td>1.008</td><td>5.2e-01</td></tr>
<tr><td>rs10888690</td><td>T (0.570)</td><td>0.979</td><td>(0.973 - 0.986)</td><td>1.2e-09</td><td>T (0.632)</td><td>0.988</td><td>2.0e-02*</td></tr>
<tr><td>rs66499095</td><td>I (0.804)</td><td>0.977</td><td>(0.970 - 0.985)</td><td>4.1e-09</td><td>I (0.766)</td><td>0.999</td><td>9.3e-01</td></tr>
<tr><td>rs2032905</td><td>G (0.577)</td><td>1.020</td><td>(1.013 - 1.026)</td><td>4.3e-09</td><td>G (0.579)</td><td>1.007</td><td>1.8e-01</td></tr>
<tr><td>rs1855263</td><td>G (0.672)</td><td>1.020</td><td>(1.013 - 1.027)</td><td>1.0e-08</td><td>G (0.675)</td><td>1.005</td><td>3.1e-01</td></tr>
<tr><td>rs35847492</td><td>G (0.707)</td><td>0.979</td><td>(0.972 - 0.986)</td><td>1.5e-08</td><td>G (0.678)</td><td>0.992</td><td>1.1e-01</td></tr>
<tr><td>rs10852372</td><td>C (0.592)</td><td>1.019</td><td>(1.012 - 1.026)</td><td>1.6e-08</td><td>C (0.600)</td><td>1.011</td><td>2.9e-02*</td></tr>
<tr><td>rs13421417</td><td>T (0.689)</td><td>0.981</td><td>(0.974 - 0.987)</td><td>1.8e-08</td><td>G (0.655)</td><td>0.993</td><td>1.8e-01</td></tr>
<tr><td>rs2920991</td><td>T (0.499)</td><td>0.982</td><td>(0.976 - 0.989)</td><td>4.8e-08</td><td>G (0.513)</td><td>0.982</td><td>6.4e-4**</td></tr>
<tr><td rowspan="2">rs12334416/ rs35840532†</td><td>C (0.532)</td><td>0.982</td><td>(0.976 - 0.988)</td><td>2.1e-08</td><td rowspan="2">I (0.553)</td><td rowspan="2">0.990</td><td rowspan="2">5.3e-02</td></tr>
<tr><td>I (0.522)</td><td>0.983</td><td>(0.977 - 0.989)</td><td>1.1e-07</td></tr>
</table>

*** denotes if the rsID has reached Bonferroni-corrected level of statistical significance (P < 0.005), * denotes if the rsID has reached nominal significance (P < 0.05), † rsID was identified to be in high LD with rs12334416 ($r^2$= 0.971), which was not included in Steinthorsdottir's summary statistics. EAF = effect allele frequency; OR = odds ratio; 95%CI = 95% confidence interval*

**Supplementary Table 5:** Intercepts and 95% CIs from MR analysis

| Exposure | Sporadic Miscarriage | | | Recurrent Miscarriage | | |
|---|---|---|---|---|---|---|
| | Intercept | 95% CI | p-value | Intercept | 95% CI | p-value |
| Ever tobacco smoker | 0.00023 | -0.00078 - 0.0012 | 0.66 | 0.00096 | -0.0011 - 0.0030 | 0.37 |
| Pack-years | 0.0027* | 0.001 - 0.0044 | 0.0018 | 0.0056* | 0.0022 - 0.0090 | 0.0016 |
| Current caffeine consumption | 0.00096 | -0.00093 - 0.0028 | 0.28 | 0.0016 | -0.0015 - 0.0047 | 0.32 |
| Heavy caffeine drinker | 0.0012 | -0.00061 - 0.0029 | 0.20 | 0.00022 | -0.0033 - 0.0038 | 0.90 |

*Symbols correspond to the P significance level; *P < 0.05. 95%CI = 95% confidence interval*

**Supplementary Table 6:** Steiger filtered random effects IVW results from Mendelian Randomization analysis

| Exposure | Outcome | SNPs | OR (95% CI) | p-value |
|---|---|---|---|---|
| Ever tobacco smoker | Sporadic miscarriage | 880 | 1.13 (1.11 - 1.15) | 5.20e-42 |
| Ever tobacco smoker | Recurrent miscarriage | 777 | 1.17 (1.13 - 1.20) | 1.72e-22 |
| Current caffeine consumption (continuous variable) | Recurrent miscarriage | 125 | 1.48 (1.09- 2.01) | 1.23e-1 |
| Heavy caffeine drinker | Recurrent miscarriage | 109 | 1.00 (0.99 - 1.11) | 1.34e-1 |

*Presented estimates are from analyses where SNPs were removed after Steiger filtering. In other words, when an MR analysis depicted in Figure 2 or 3 is not presented above, it means that no SNPs were removed following Steiger filtering in that MR analysis. OR = odds ratio; 95%CI = 95% confidence interval*

**Supplementary Table 7:** Variant-to-gene mapping (LD $r^2$ > 0.8 with coding, eQTL, or pQTL variants) for sporadic miscarriage per population

| Lead variant | Coding gene (variant) | pQTL (variant) | eQTL (variant) | Ancestry |
|---|---|---|---|---|
| rs66499095 | - | - | *MRPS21* (rs72698868[57], rs12729727[58], rs4970946[58], rs12116970[58,59], rs16836940[60], rs1050818[58,61], rs71652043[61], rs7542510[62], rs112088857[63]) | European, African American, Latinx |
| | | | *RPRD2* (rs10888581[65], rs6670974[62], rs34353365[58], rs12139488[65], rs36038795*, rs6694168[58], rs35699789[58], rs12116970[58,64], rs68097764[66], rs16836940[60,66], rs34391279[67]) | European, African American, Latinx |
| | | | *TARS2* (rs12116970[65]) | European |
| | | *ECM1* (rs11582094[70]) | - | African American |
| rs12334416 | - | - | *SOX7* (rs1124010 [65], rs6601521 [65]) | European, Latinx |
| | | | *GATA4* (rs2409672[68]) | European, Latinx |
| | | | *RP1L1* (rs6601530*) | European |
| rs2032905 | - | - | *TSHZ3* (rs2032905[69]) | European, African American, Latinx |
| rs13107325 | *SLC39A8* (rs13107325) | - | - | African American |

**Meta-analysis done by 23andMe (see Supplementary Table 12)*

**Supplementary Table 8: Effect allele frequencies (EAF) for sporadic miscarriage top hits per ancestry group**

| rsID | Effect Allele / Non-Effect Allele | EAF in Controls [European/ Latinx/ African American] | EAF in Cases [European/ Latinx/ African American] |
|---|---|---|---|
| rs13107325 | T / C | 0.081/ 0.061/ 0.024 | 0.083/ 0.063/ 0.025 |
| rs10888690 | T / C | 0.59/ 0.63/ 0.46 | 0.58/ 0.63/ 0.46 |
| rs66499095:CA_C | I / D | 0.78/ 0.84/ 0.70 | 0.78/ 0.83/ 0.70 |
| rs2032905 | G / A | 0.60/ 0.57/ 0.38 | 0.61/ 0.57/ 0.38 |
| rs1855263 | G / A | 0.67/ 0.67/ 0.60 | 0.67/ 0.68/ 0.61 |
| rs35847492 | G / C | 0.68/ 0.74/ 0.81 | 0.68/ 0.74/ 0.80 |
| rs10852372 | C / A | 0.59/ 0.56/ 0.84 | 0.60/ 0.57/ 0.85 |
| rs13421417 | T / A | 0.66/ 0.73/ 0.89 | 0.66/ 0.73/ 0.8 |
| rs12334416 | C / A | 0.54/ 0.50/ 0.66 | 0.54/ 0.50/ 0.65 |
| rs2920991 | T / A | 0.50/ 0.49/ 0.73 | 0.50/ 0.49/ 0.73 |

**Supplementary Table 9: Sample overlap for each exposure and outcome for Mendelian randomization analysis**

| Exposure name (N) | Outcome name (N) | Sample of exposure and not in outcome (N) | Sample overlap of exposure and outcome (N) | Sample of outcome and not in exposure (N) |
|---|---|---|---|---|
| Ever tobacco user (3,609,906) | Sporadic Miscarriage (715,286) | 3,179,684 | 430,222 | 285,064 |
| | Recurrent Miscarriage (509,688) | 3,308,504 | 301,402 | 208,286 |
| Pack-years (593,494) | Sporadic Miscarriage (715,286) | 421,367 | 172,127 | 543,159 |
| | Recurrent Miscarriage (509,688) | 473,762 | 119,732 | 389,956 |
| Current caffeine consumption (854,473) | Sporadic Miscarriage (715,286) | 552,190 | 302,283 | 413,003 |
| | Recurrent Miscarriage (509,688) | 640,098 | 214,375 | 295,313 |
| Heavy caffeine drinker (912,999) | Sporadic Miscarriage (715,286) | 594,141 | 318,858 | 396,428 |
| | Recurrent Miscarriage (509,688) | 687,336 | 225,663 | 284,025 |

**Supplementary Table 10: Rerun of European sporadic miscarriage GWAS Top Hits, using age at first pregnancy as a covariate**

| rsID | Main Analysis (adjusted for age at survey)<br>Cases = 284,795<br>Controls = 430,491 | Main Analysis (adjusted for age at survey and restricted to individuals with available age at first pregnancy information)<br>Cases = 189,445<br>Controls = 260,169 | Sensitivity Analysis (adjusted for age at first pregnancy)<br>Cases = 189,445<br>Controls= 260,169 |
|---|---|---|---|
| | **OR** | **OR** | **OR** |
| rs10888690 | 0.98 | 0.98 | 0.98 |
| rs71086513 | 1.03 | 1.02 | 1.02 |
| rs13421417 | 0.98 | 0.98 | 0.98 |
| rs775261971 | 0.43 | 0.43 | 0.44 |
| rs35225200 | 1.04 | 1.04 | 1.04 |
| rs17360811 | 1.02 | 1.03 | 1.02 |
| rs2920991 | 0.98 | 0.98 | 0.99 |
| rs1962430 | 1.02 | 1.02 | 1.02 |
| rs773587903 | 2.09 | 2.05 | 2.07 |

*95%CI = 95% confidence interval*

**Supplementary Table 11: Intercepts and 95% CIs from split-sample MR analysis**

| Exposure | Sporadic Miscarriage | | | Recurrent Miscarriage | | |
|---|---|---|---|---|---|---|
| | Intercept | 95% CI | p-value | Intercept | 95% CI | p-value |
| Ever tobacco smoker | -0.00014 | (-0.0013 - 0.00099) | 0.81 | -0.00035 | (-0.0027 - 0.0020) | 0.77 |
| Pack-years | 0.0032 | (-0.00026 - 0.0067) | 0.076 | 0.0075 | (0.00061 - 0.014) | 0.038* |
| Current caffeine consumption | -0.0011 | (-0.0042 - 0.0020) | 0.50 | 0.00085 | (-0.0054 - 0.0071) | 0.79 |
| Heavy caffeine drinker | -2.60e-05 | (-0.0049 - 0.0049) | 0.99 | 0.00057 | (-0.0072 - 0.0083) | 0.89 |

*Symbols correspond to the P significance level; *P < 0.05. 95%CI = 95% confidence interval*

#### Supplementary Table 12: Public datasets that are internally processed using 23andMe eQTL pipelines

| 23andMe eQTL pipeline | Title | Tissue | Ethnicity | Sample |
|---|---|---|---|---|
| First pipeline | TTAM2_AFR_LCL* | Lymphoblast | African | 659 |
| Second pipeline | TTAM_BLOOD_META** | Venous blood | Multi-ethnic | 2,712 |

**based on an external study sample set with data generated in-house, **consisted of three studies: PPMI (Craig, D. W. et al. RNA sequencing of whole blood reveals early alterations in immune cells and gene expression in Parkinson's disease. Nature aging 1, (2021).), GTEx v8 (The GTEx Consortium atlas of genetic regulatory effects across human tissues. Science 369, (2020).), and BRGR (based on 23andMe customer data)*

**Supplementary Figure 1: Study flowchart depicting how miscarriage cases and controls were enrolled into study**

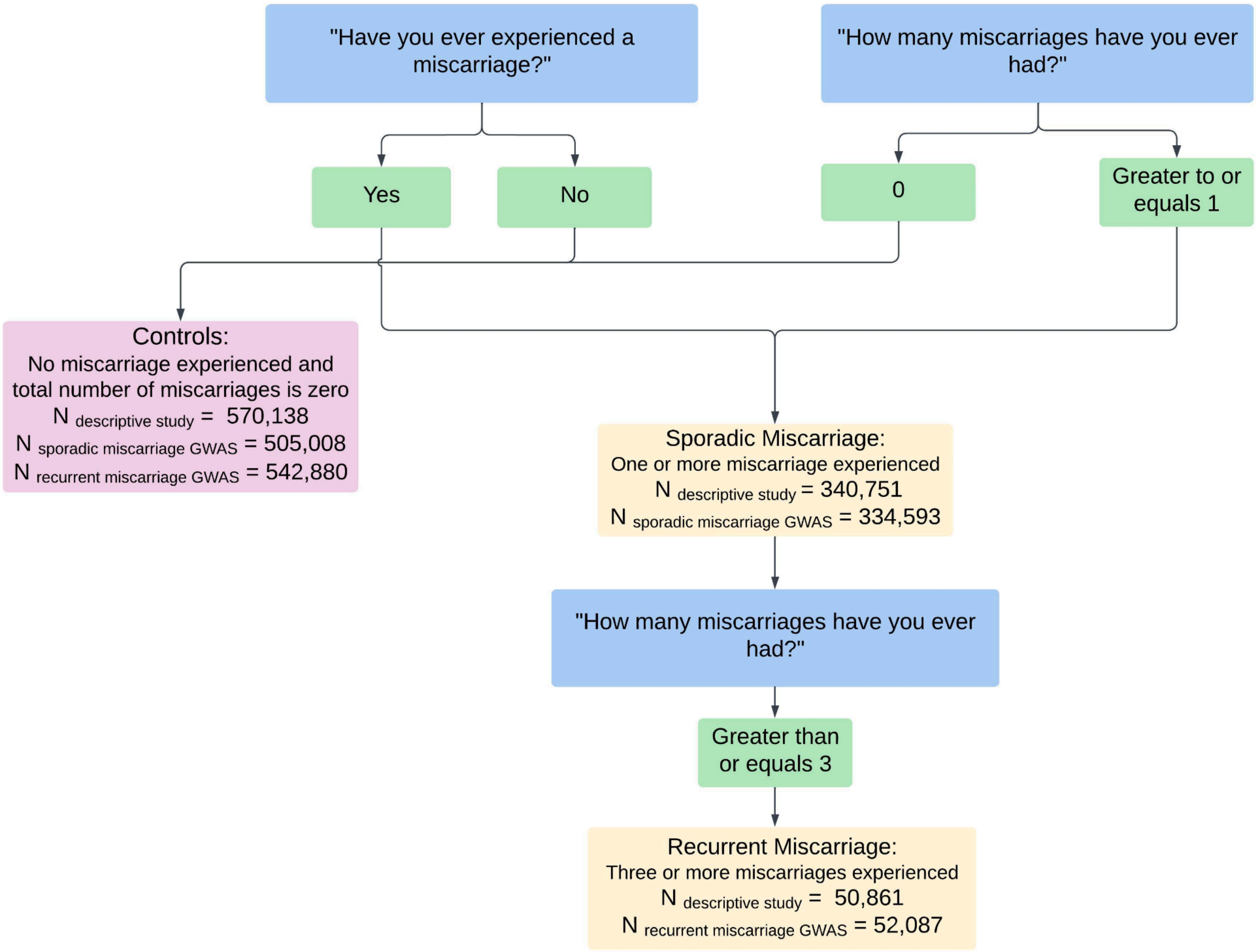

**Supplementary Figure 2: Manhattan plots for both recurrent and sporadic miscarriage variables and each population prior to trans-ancestral meta-analysis**

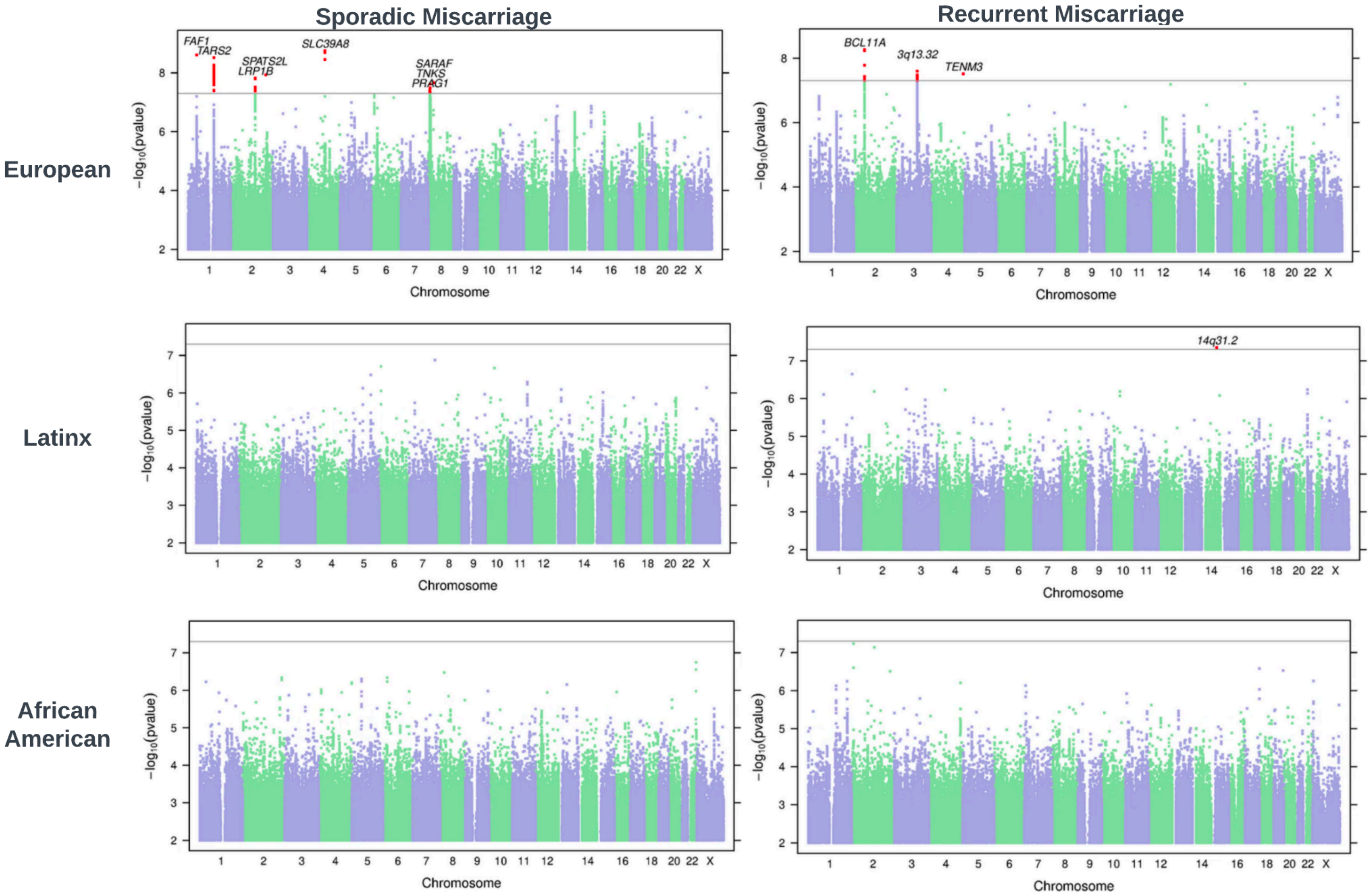


*The nearest gene to each index SNP is indicated above each association peak. SNPs achieving genome-wide significance are highlighted in red.*

**Supplementary Figure 3: Locus zoom plots for SNPs that reached genome-wide significance on trans-ancestral analysis of (a) sporadic miscarriage and (b) recurrent miscarriage**

a

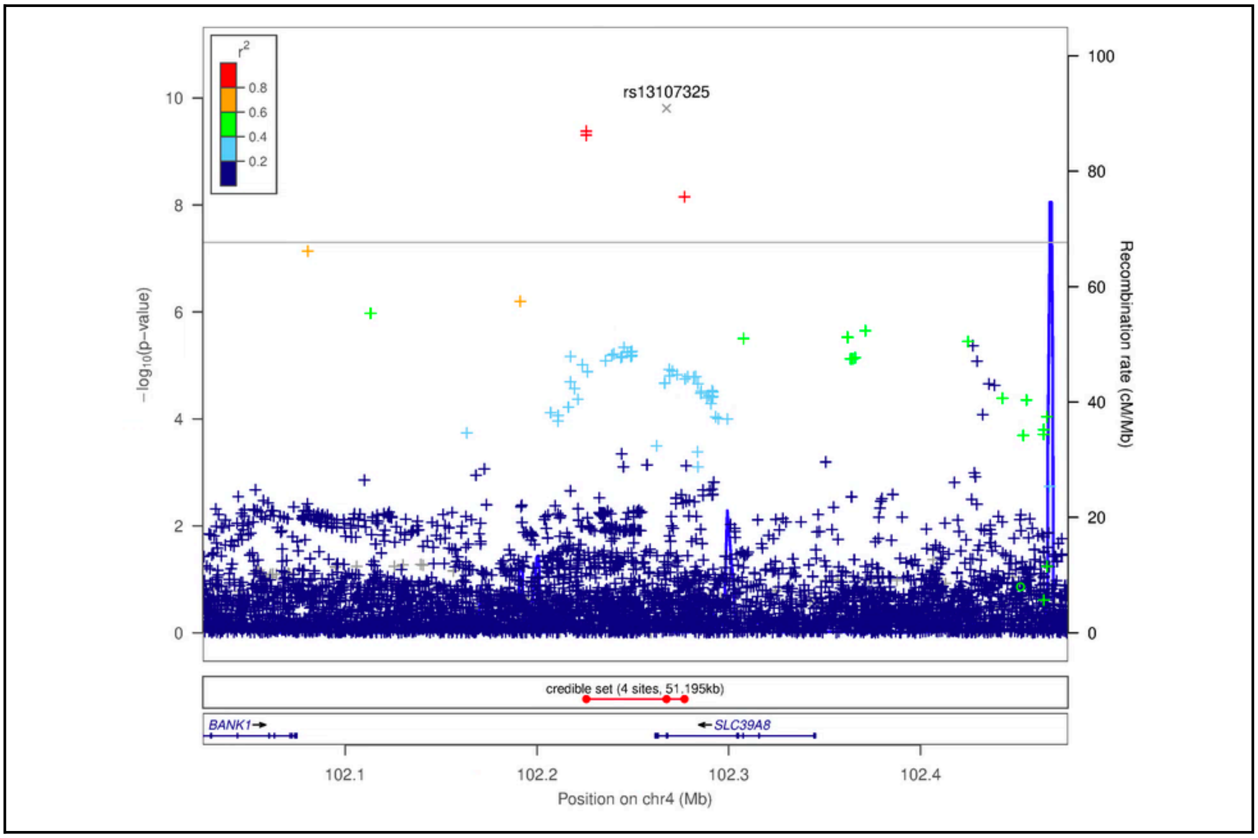

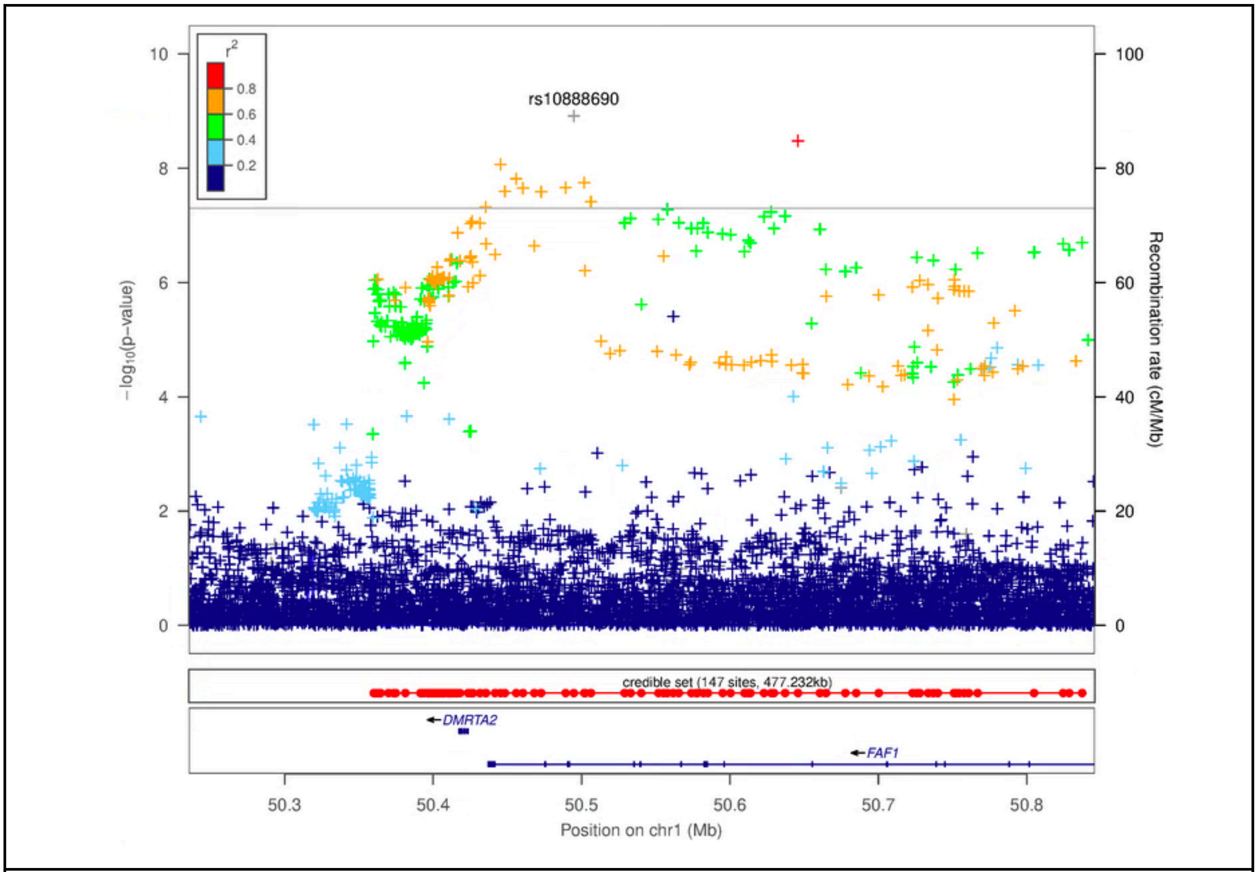
rs10888690
credible set (147 sites, 477.232kb)
DMRTA2
FAF1
Position on chr1 (Mb)
Recombination rate (cM/Mb)

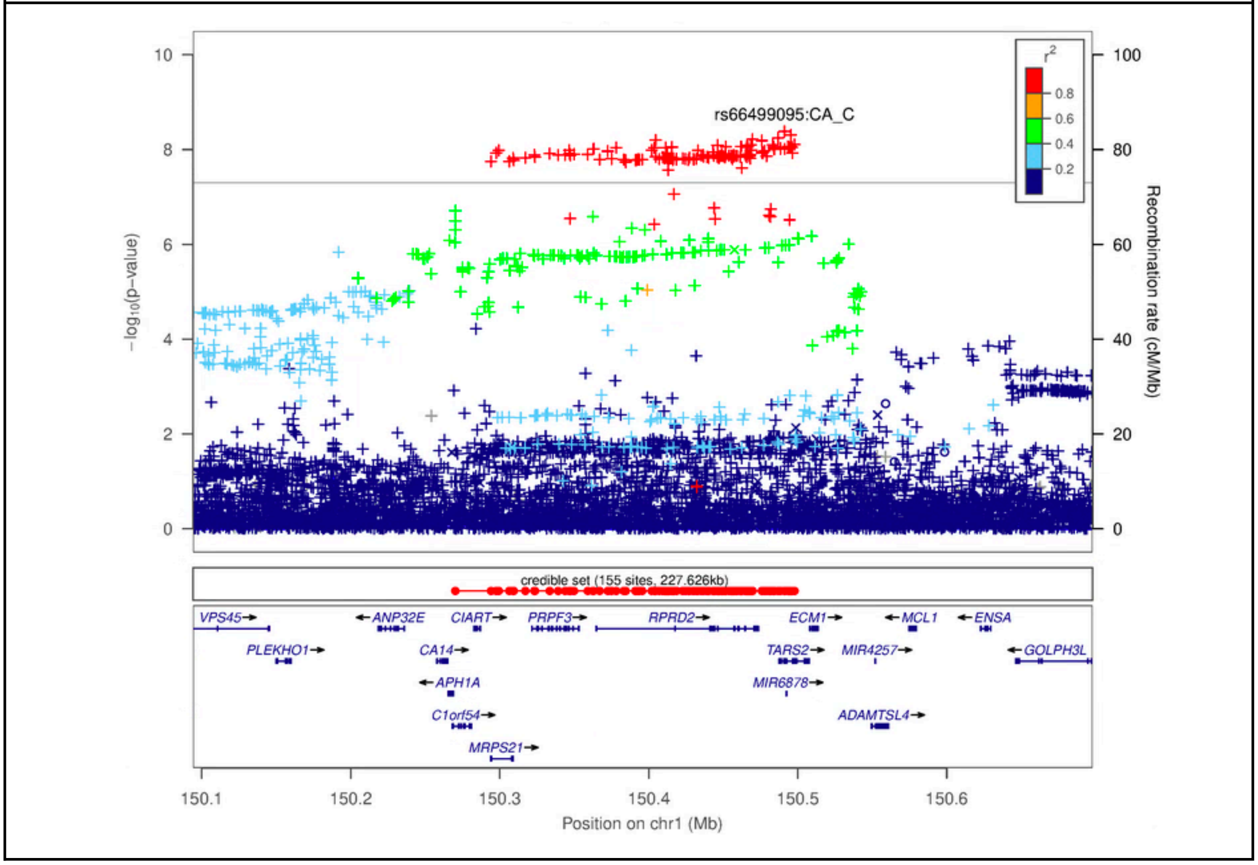
rs66499095:CA_C
credible set (155 sites, 227.626kb)
VPS45
ANP32E
CIART
PRPF3
RPRD2
ECM1
MCL1
ENSA
PLEKHO1
CA14
TARS2
MIR4257
GOLPH3L
APH1A
MIR6878
C1orf54
ADAMTSL4
MRPS21
Position on chr1 (Mb)
Recombination rate (cM/Mb)

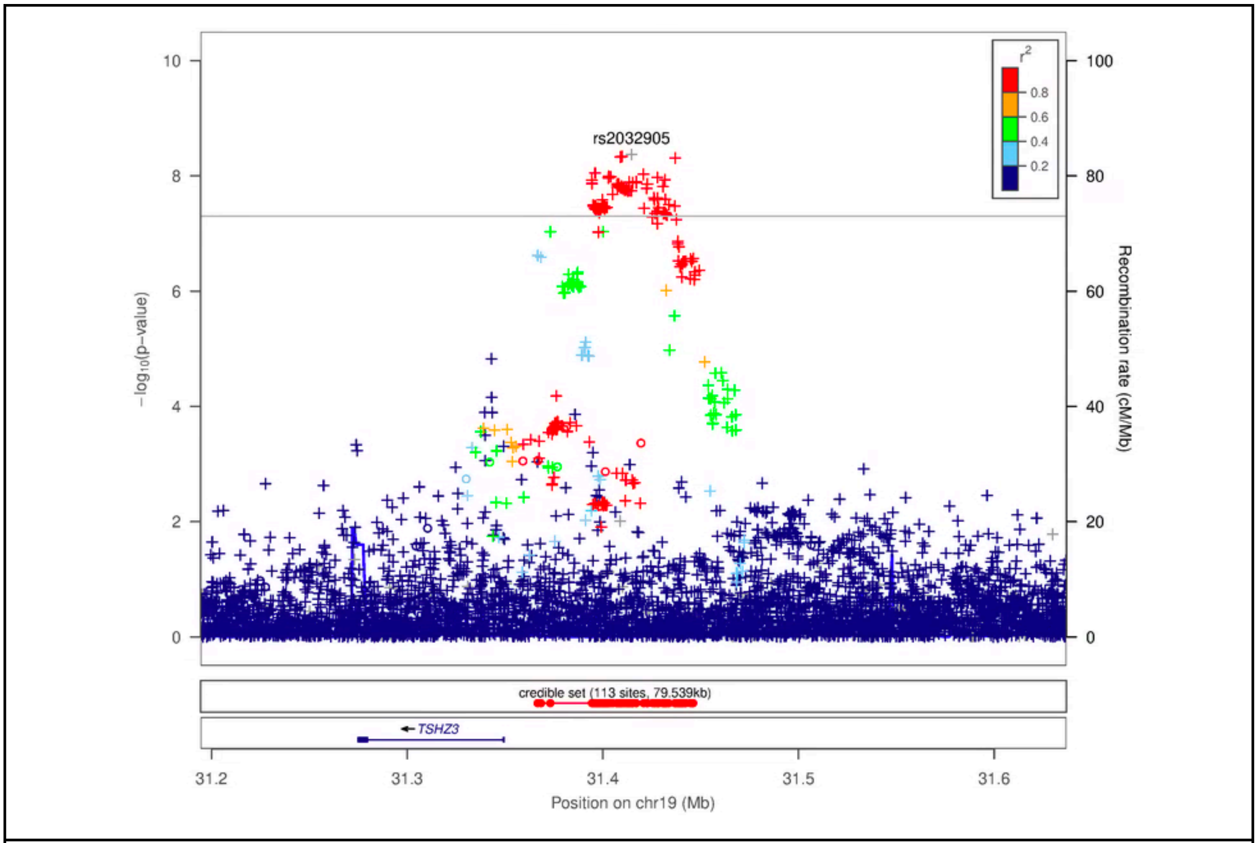
rs2032905
r2
0.8
0.6
0.4
0.2
−log10(p−value)
Recombination rate (cM/Mb)
credible set (113 sites, 79.539kb)
TSHZ3
31.2
31.3
31.4
31.5
31.6
Position on chr19 (Mb)

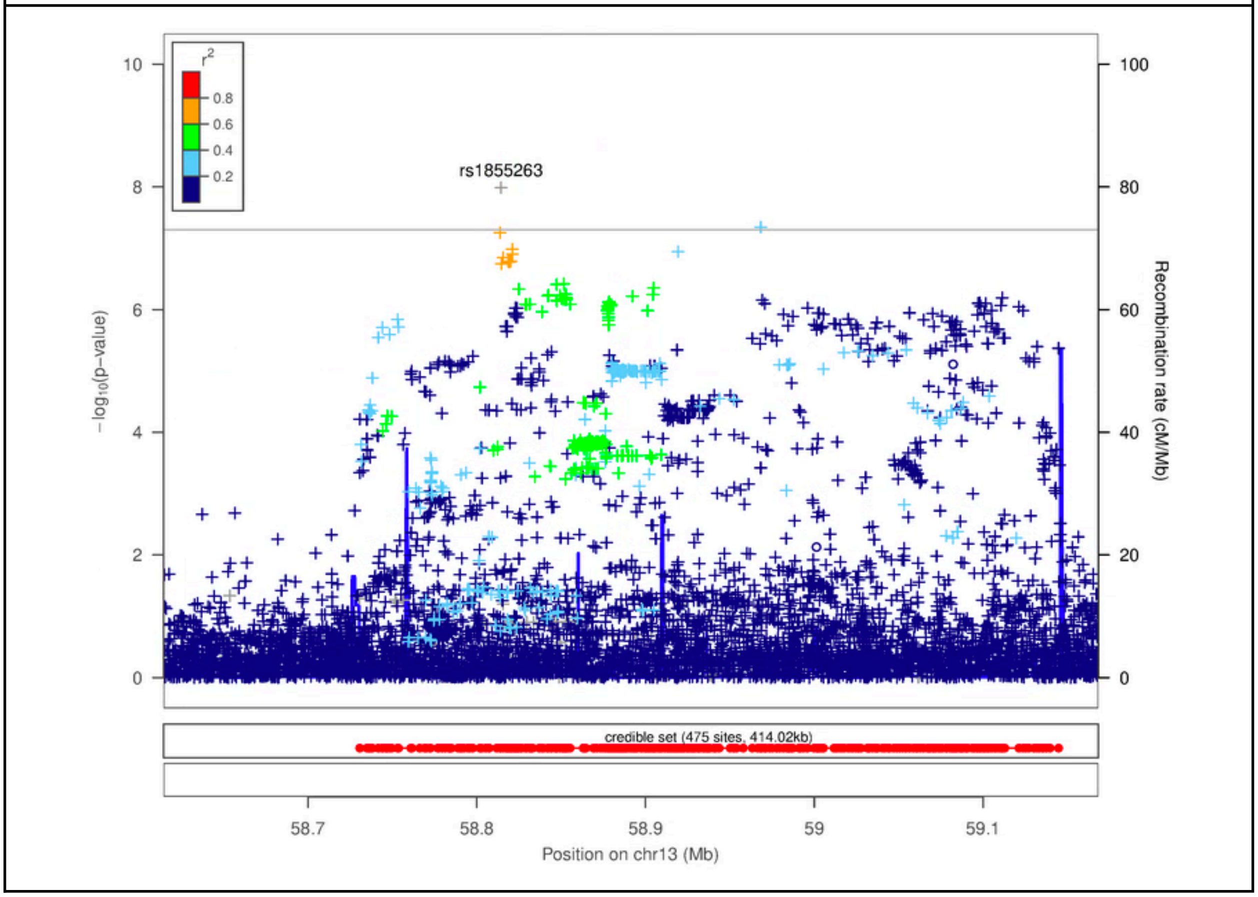
rs1855263
r2
0.8
0.6
0.4
0.2
−log10(p−value)
Recombination rate (cM/Mb)
credible set (475 sites, 414.02kb)
58.7
58.8
58.9
59
59.1
Position on chr13 (Mb)

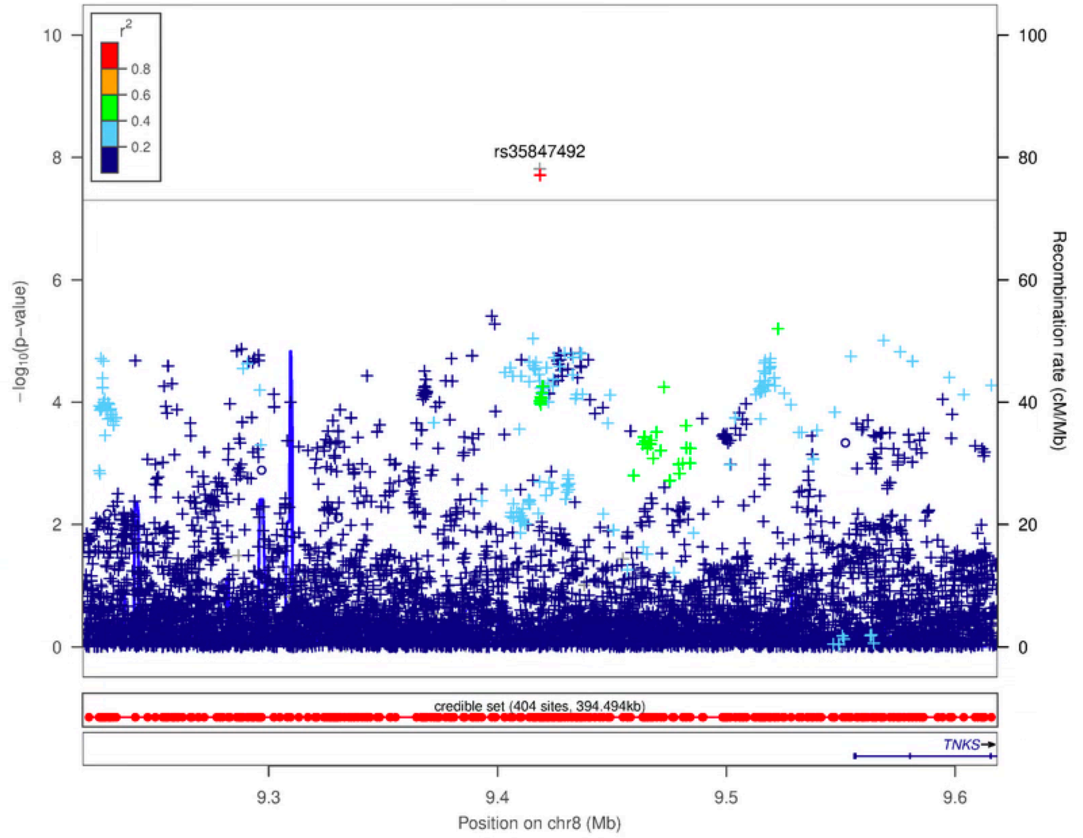

r²
0.8
0.6
0.4
0.2
rs35847492
−log10(p−value)
Recombination rate (cM/Mb)
credible set (404 sites, 394.494kb)
TNKS
Position on chr8 (Mb)


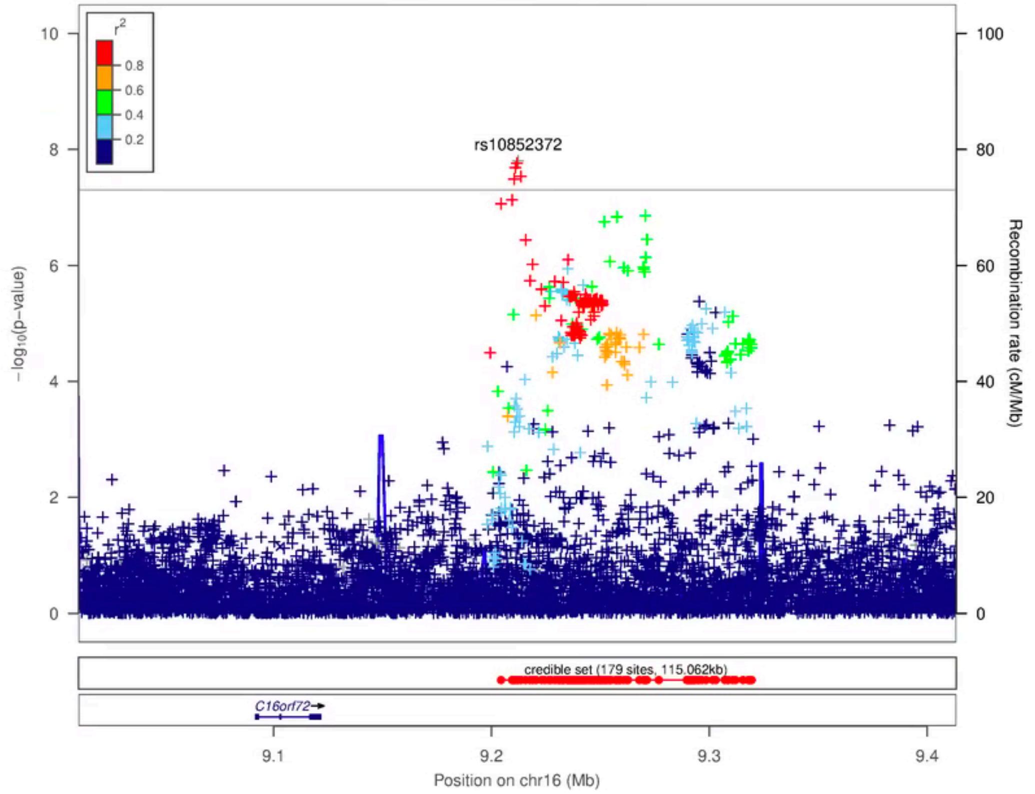

r²
0.8
0.6
0.4
0.2
rs10852372
−log10(p−value)
Recombination rate (cM/Mb)
credible set (179 sites, 115.062kb)
C16orf72
Position on chr16 (Mb)

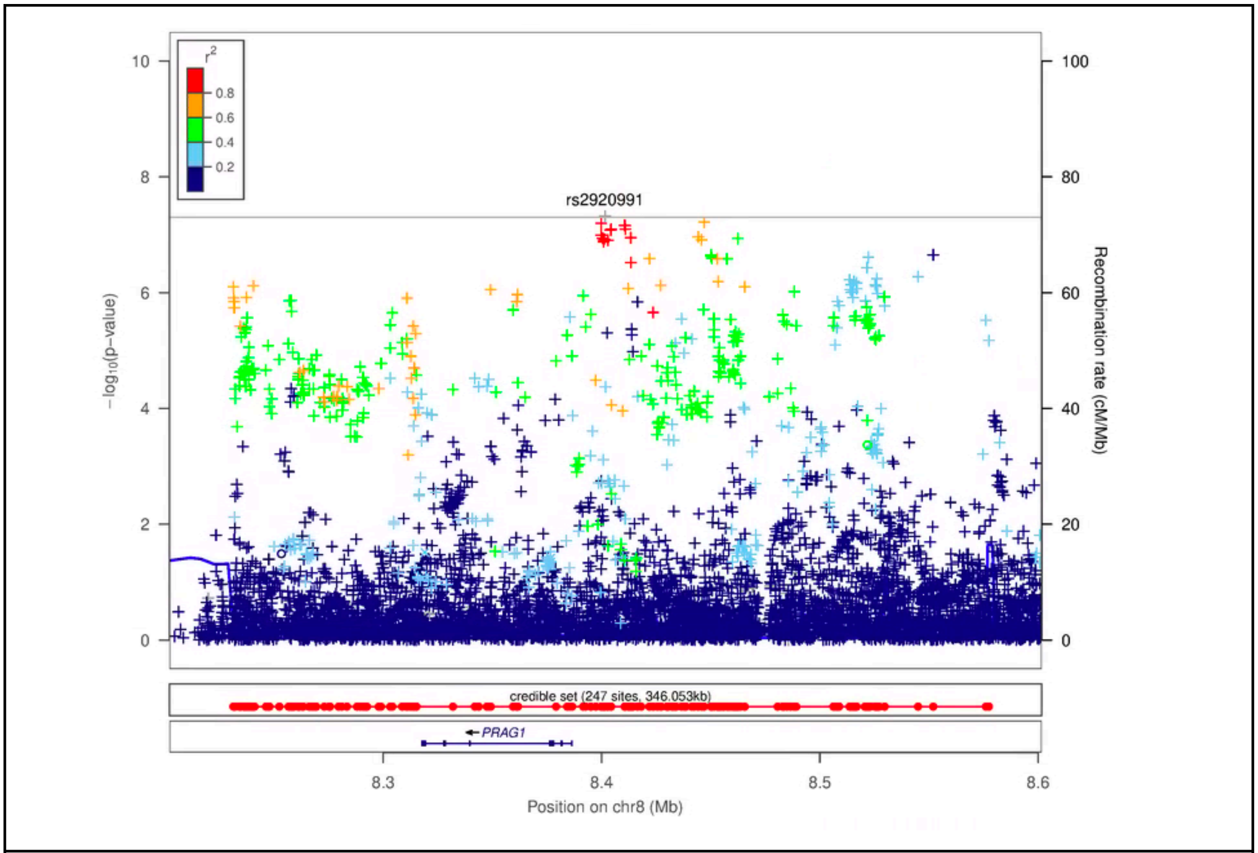
r2
0.8
0.6
0.4
0.2
rs2920991
−log10(p−value)
Recombination rate (cM/Mb)
credible set (247 sites, 346.053kb)
PRAG1
8.3
8.4
8.5
8.6
Position on chr8 (Mb)

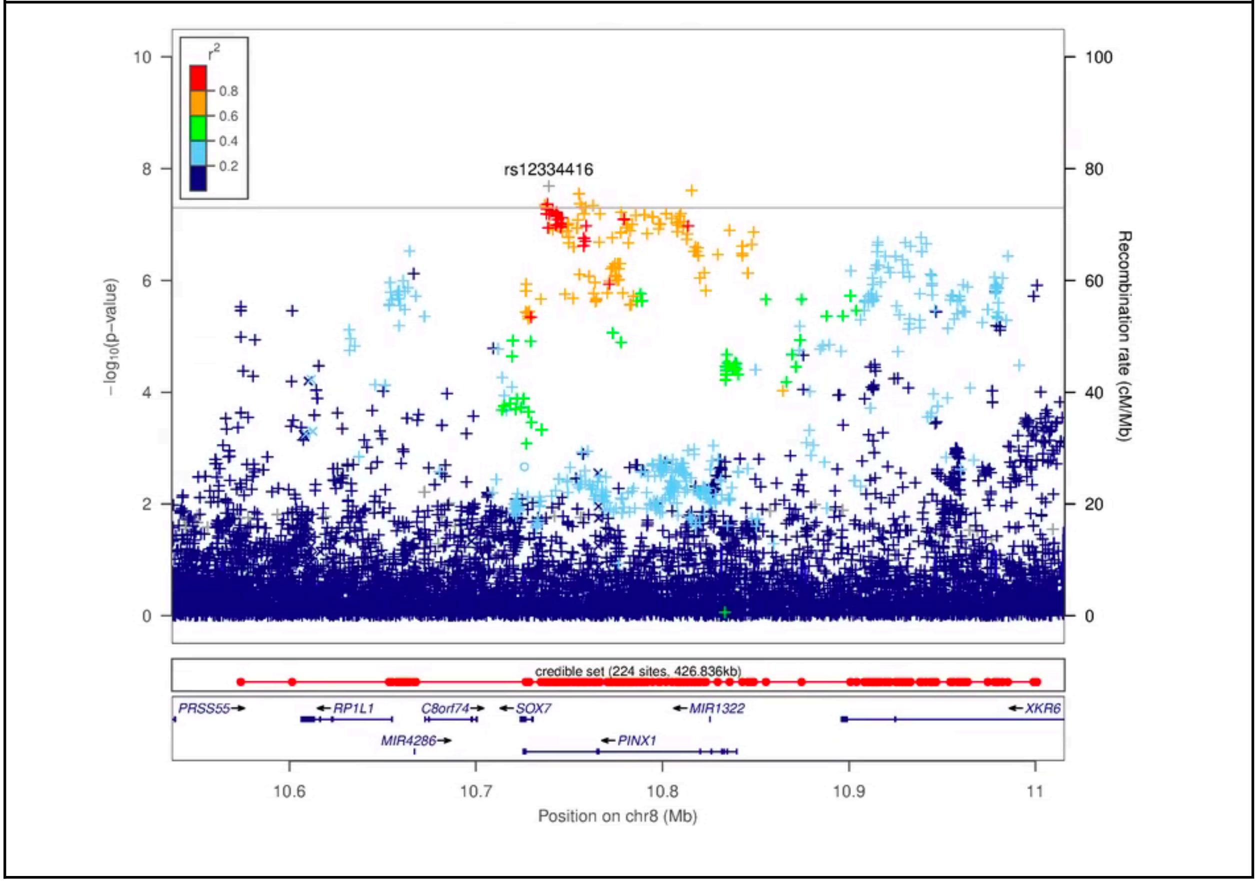
r2
0.8
0.6
0.4
0.2
rs12334416
−log10(p−value)
Recombination rate (cM/Mb)
credible set (224 sites, 426.836kb)
PRSS55
RP1L1
C8orf74
SOX7
MIR1322
XKR6
MIR4286
PINX1
10.6
10.7
10.8
10.9
11
Position on chr8 (Mb)

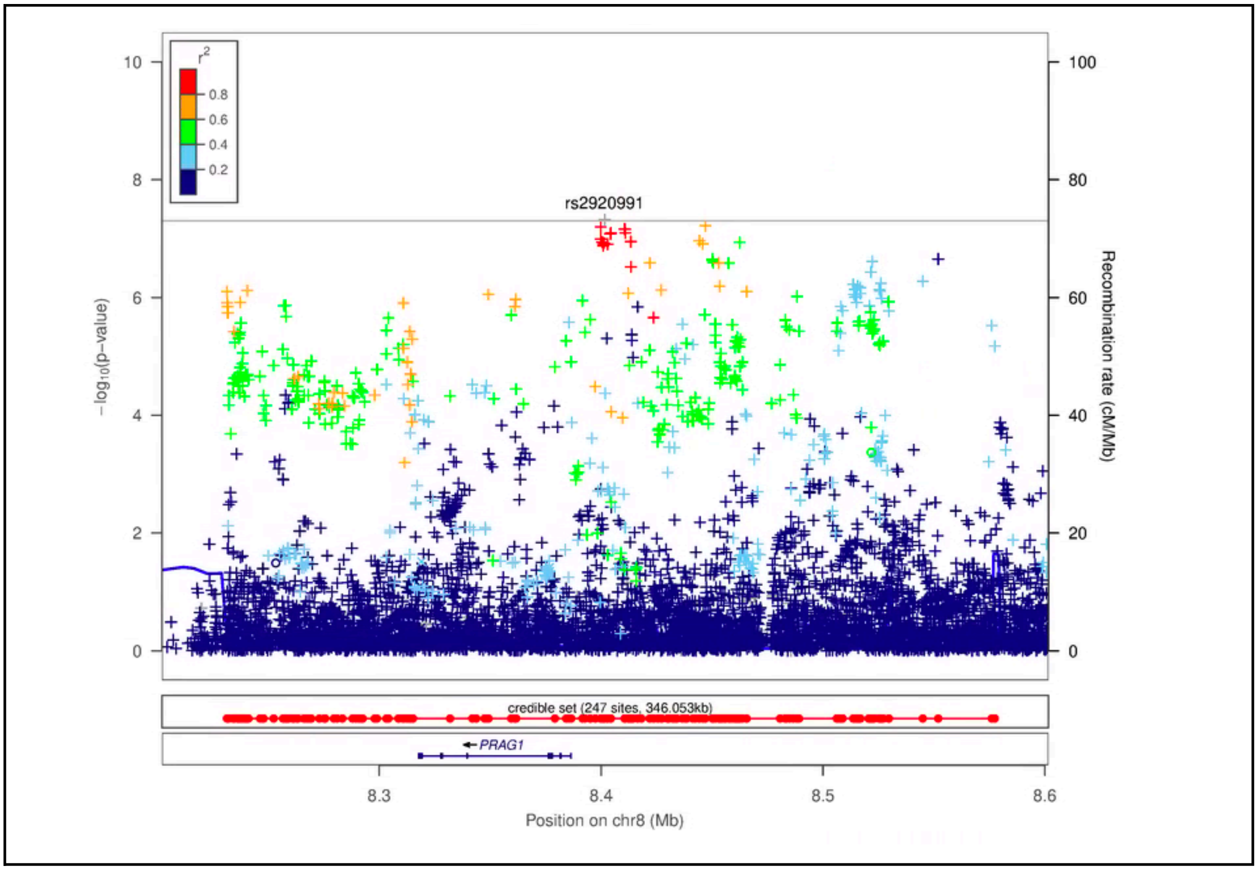
rs2920991
-log10(p-value)
Recombination rate (cM/Mb)
credible set (247 sites, 346.053kb)
PRAG1
Position on chr8 (Mb)

b

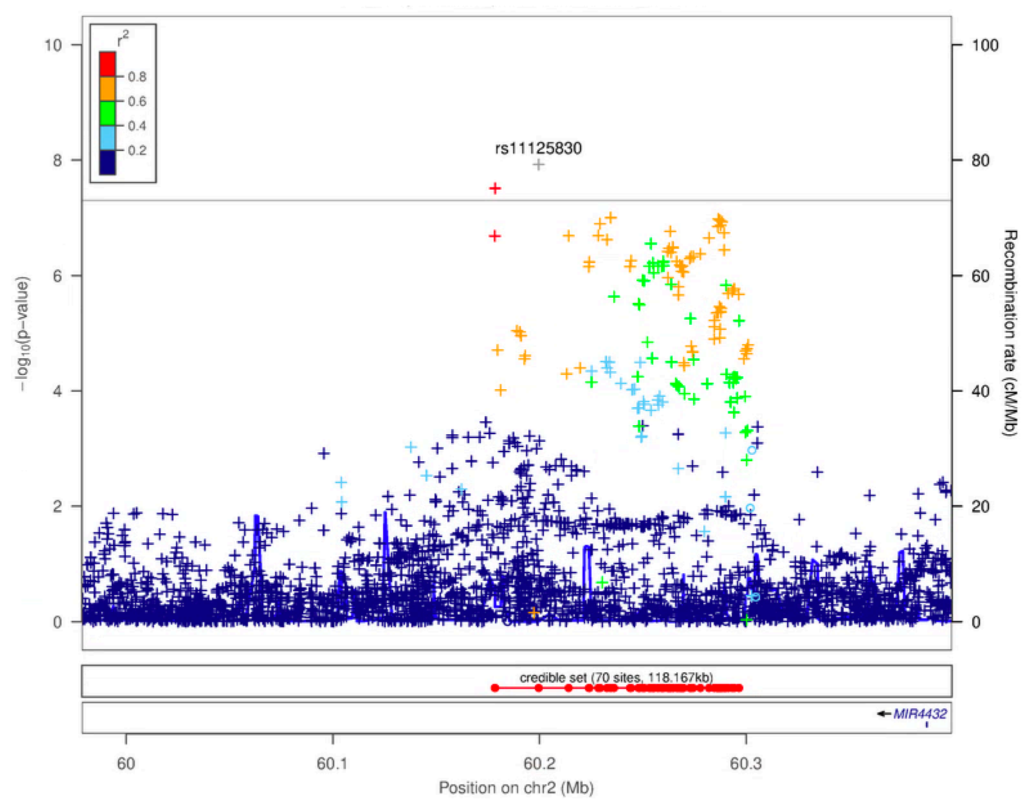
rs11125830
-log10(p-value)
Recombination rate (cM/Mb)
credible set (70 sites, 118.167kb)
MIR4432
Position on chr2 (Mb)

**Supplementary Figure 4: Forest plots of SNPs reaching GWAS significance for miscarriage comparing main analysis to sensitivity analysis (restricting cases and controls to having previously reported a pregnancy)**

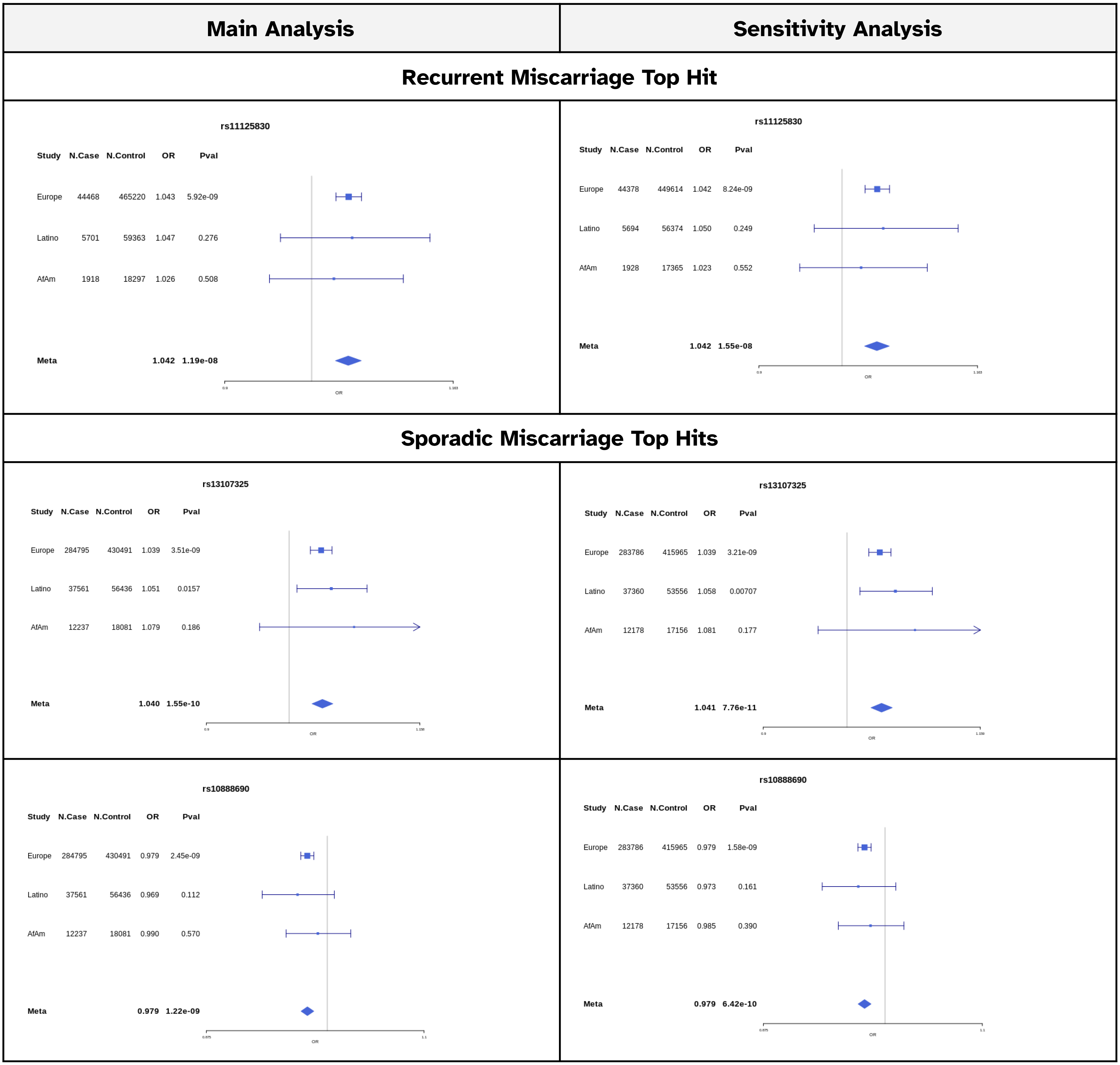

**rs66499095:CA_C**

| Study | N.Case | N.Control | OR | Pval |
|---|---|---|---|---|
| Europe | 284795 | 430491 | 0.976 | 5.95e-09 |
| Latino | 37561 | 56436 | 0.974 | 0.0490 |
| AfAm | 12237 | 18081 | 1.006 | 0.759 |
| **Meta** | | | **0.977** | **4.10e-09** |

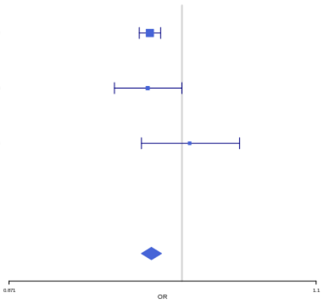


**rs66499095:CA_C**

| Study | N.Case | N.Control | OR | Pval |
|---|---|---|---|---|
| Europe | 283786 | 415965 | 0.976 | 4.13e-09 |
| Latino | 37360 | 53556 | 0.975 | 0.0607 |
| AfAm | 12178 | 17156 | 1.002 | 0.906 |
| **Meta** | | | **0.977** | **2.56e-09** |

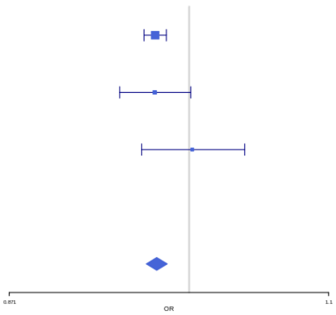


**rs2032905**

| Study | N.Case | N.Control | OR | Pval |
|---|---|---|---|---|
| Europe | 284795 | 430491 | 1.018 | 5.31e-07 |
| Latino | 37561 | 56436 | 1.036 | 0.000401 |
| AfAm | 12237 | 18081 | 1.012 | 0.492 |
| **Meta** | | | **1.020** | **4.25e-09** |

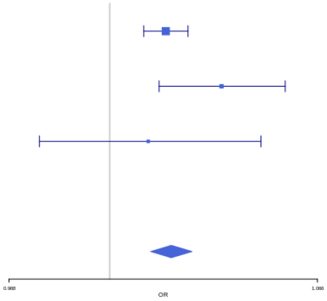


**rs2032905**

| Study | N.Case | N.Control | OR | Pval |
|---|---|---|---|---|
| Europe | 283786 | 415965 | 1.019 | 7.53e-08 |
| Latino | 37360 | 53556 | 1.037 | 0.000259 |
| AfAm | 12178 | 17156 | 1.016 | 0.382 |
| **Meta** | | | **1.021** | **3.28e-10** |

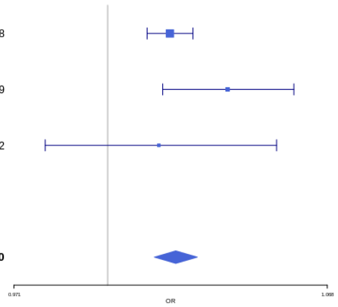


**rs1855263**

| Study | N.Case | N.Control | OR | Pval |
|---|---|---|---|---|
| Europe | 284795 | 430491 | 1.019 | 2.94e-07 |
| Latino | 37561 | 56436 | 1.023 | 0.0272 |
| AfAm | 12237 | 18081 | 1.030 | 0.0945 |
| **Meta** | | | **1.020** | **1.03e-08** |

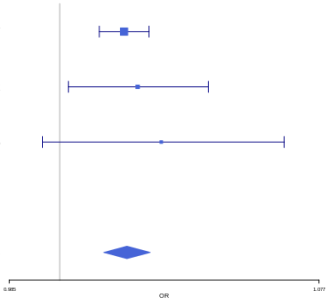


**rs1855263**

| Study | N.Case | N.Control | OR | Pval |
|---|---|---|---|---|
| Europe | 283786 | 415965 | 1.018 | 8.42e-07 |
| Latino | 37360 | 53556 | 1.025 | 0.0201 |
| AfAm | 12178 | 17156 | 1.032 | 0.0848 |
| **Meta** | | | **1.020** | **2.21e-08** |

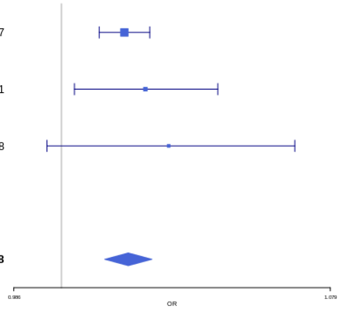


**rs35847492**

| Study | N.Case | N.Control | OR | Pval |
|---|---|---|---|---|
| Europe | 284795 | 430491 | 0.980 | 4.61e-08 |
| AfAm | 12237 | 18081 | 0.959 | 0.0513 |
| **Meta** | | | **0.979** | **1.54e-08** |

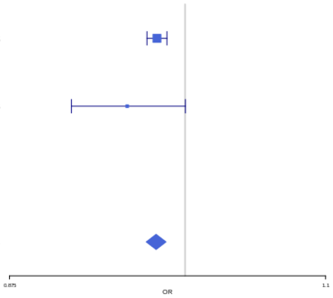


**rs35847492**

| Study | N.Case | N.Control | OR | Pval |
|---|---|---|---|---|
| Europe | 283786 | 415965 | 0.980 | 4.73e-08 |
| AfAm | 12178 | 17156 | 0.955 | 0.0373 |
| **Meta** | | | **0.979** | **1.33e-08** |

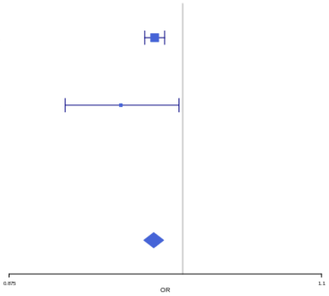

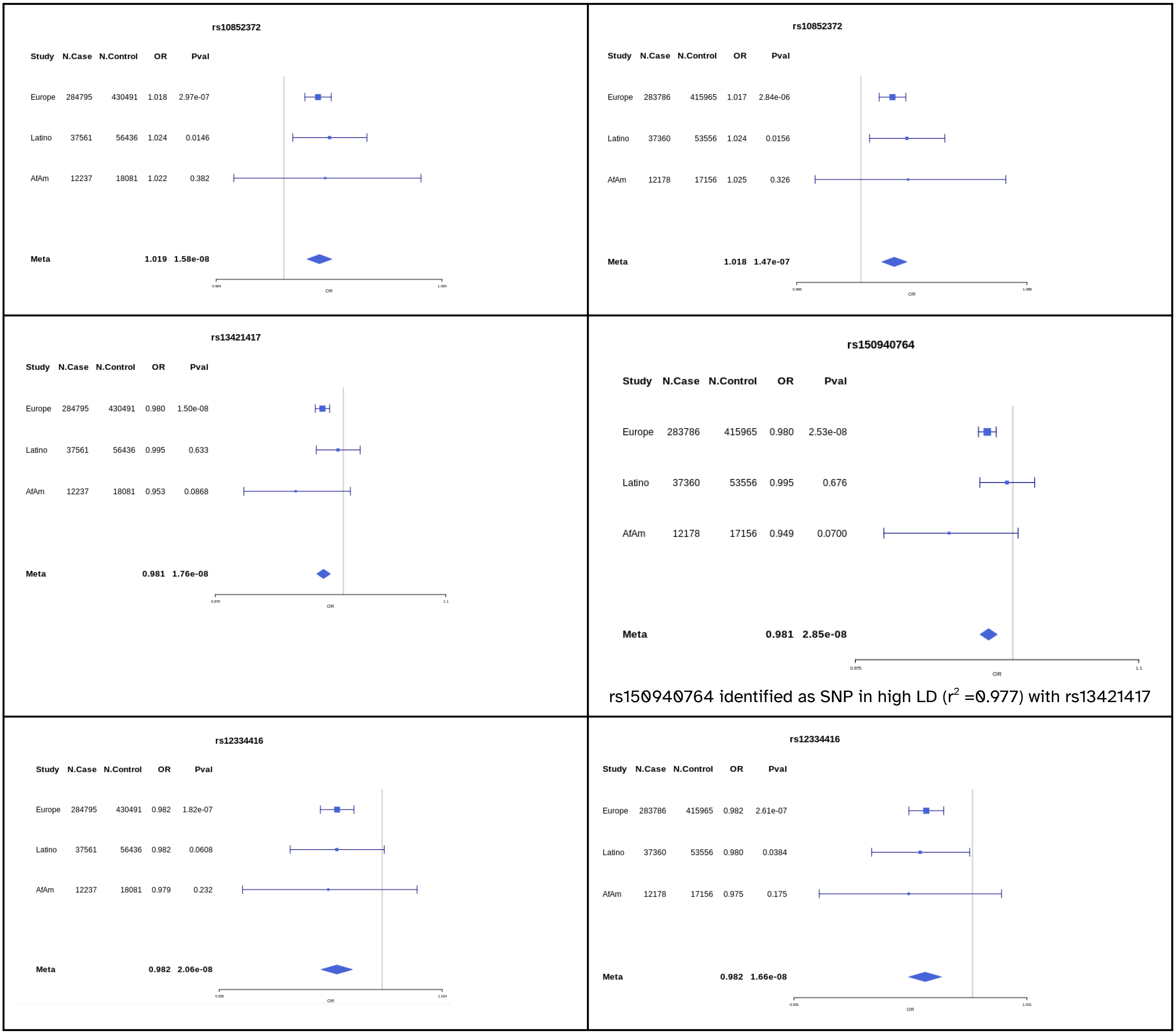
rs10852372
Study N.Case N.Control OR Pval
Europe 284795 430491 1.018 2.97e-07
Latino 37561 56436 1.024 0.0146
AfAm 12237 18081 1.022 0.382
Meta 1.019 1.58e-08
OR
rs10852372
Study N.Case N.Control OR Pval
Europe 283786 415965 1.017 2.84e-06
Latino 37360 53556 1.024 0.0156
AfAm 12178 17156 1.025 0.326
Meta 1.018 1.47e-07
OR
rs13421417
Study N.Case N.Control OR Pval
Europe 284795 430491 0.980 1.50e-08
Latino 37561 56436 0.995 0.633
AfAm 12237 18081 0.953 0.0868
Meta 0.981 1.76e-08
OR
rs150940764
Study N.Case N.Control OR Pval
Europe 283786 415965 0.980 2.53e-08
Latino 37360 53556 0.995 0.676
AfAm 12178 17156 0.949 0.0700
Meta 0.981 2.85e-08
0.875
1.1
OR
rs150940764 identified as SNP in high LD (r2 =0.977) with rs13421417
rs12334416
Study N.Case N.Control OR Pval
Europe 284795 430491 0.982 1.82e-07
Latino 37561 56436 0.982 0.0608
AfAm 12237 18081 0.979 0.232
Meta 0.982 2.06e-08
OR
rs12334416
Study N.Case N.Control OR Pval
Europe 283786 415965 0.982 2.61e-07
Latino 37360 53556 0.980 0.0384
AfAm 12178 17156 0.975 0.175
Meta 0.982 1.66e-08
OR

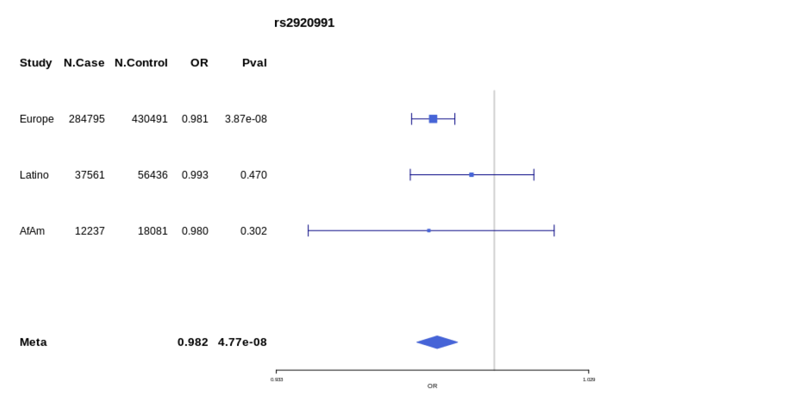
rs2920991
Study N.Case N.Control OR Pval
Europe 284795 430491 0.981 3.87e-08
Latino 37561 56436 0.993 0.470
AfAm 12237 18081 0.980 0.302
Meta 0.982 4.77e-08
OR

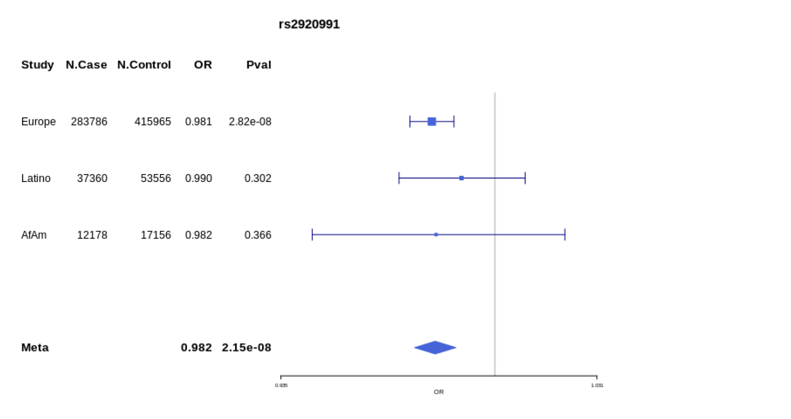
rs2920991
Study N.Case N.Control OR Pval
Europe 283786 415965 0.981 2.82e-08
Latino 37360 53556 0.990 0.302
AfAm 12178 17156 0.982 0.366
Meta 0.982 2.15e-08
OR

### Supplementary Figure 5: Manhattan plots of sensitivity analysis

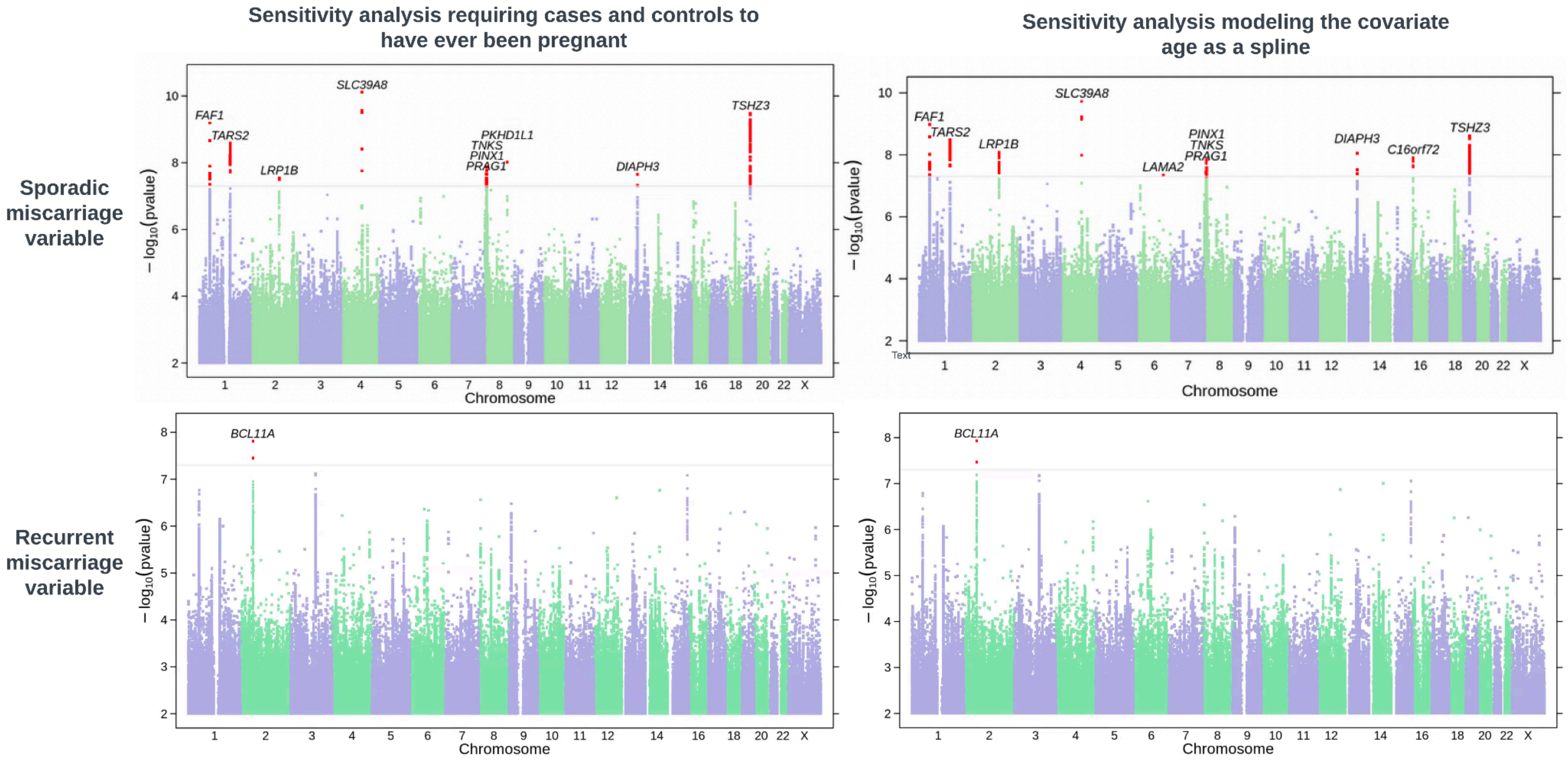


*The nearest gene to each index SNP is indicated above each association peak. SNPs achieving genome-wide significance are highlighted in red.*

#### Supplementary Figure 6: MR Scatter plots

| Sporadic miscarriage | Recurrent miscarriage |
|---|---|
| **Tobacco variables** | |

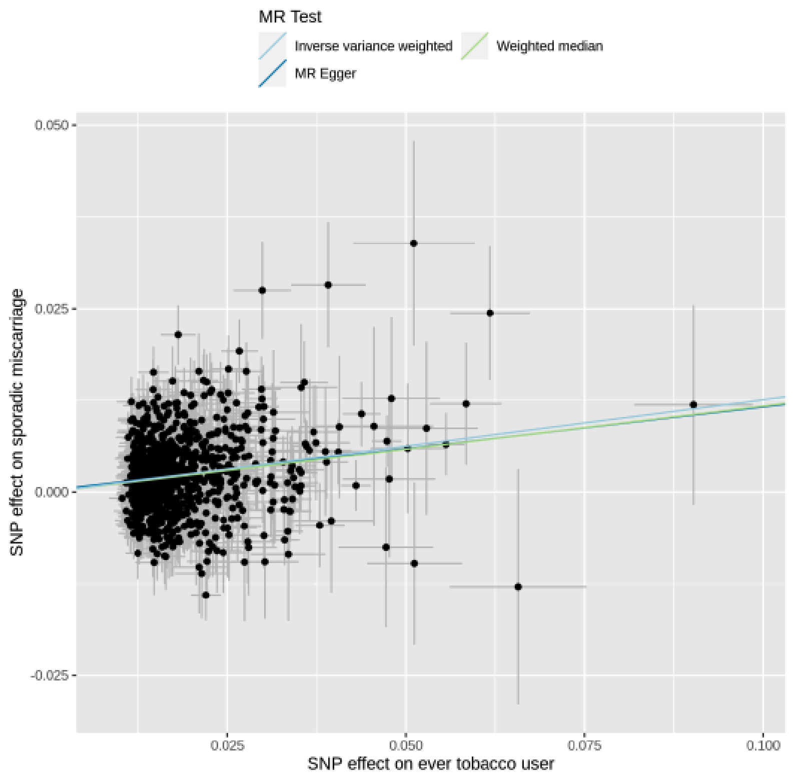


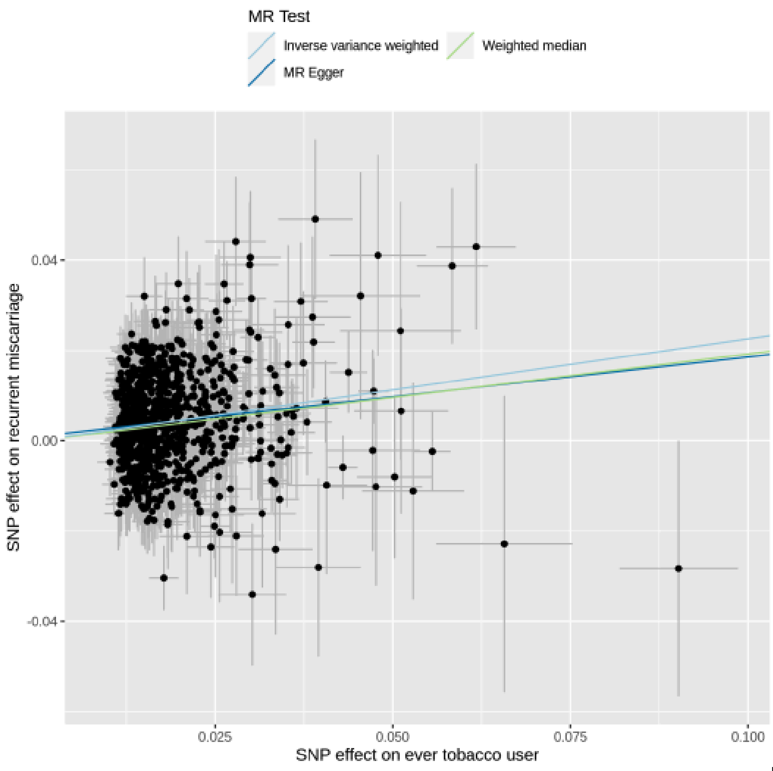

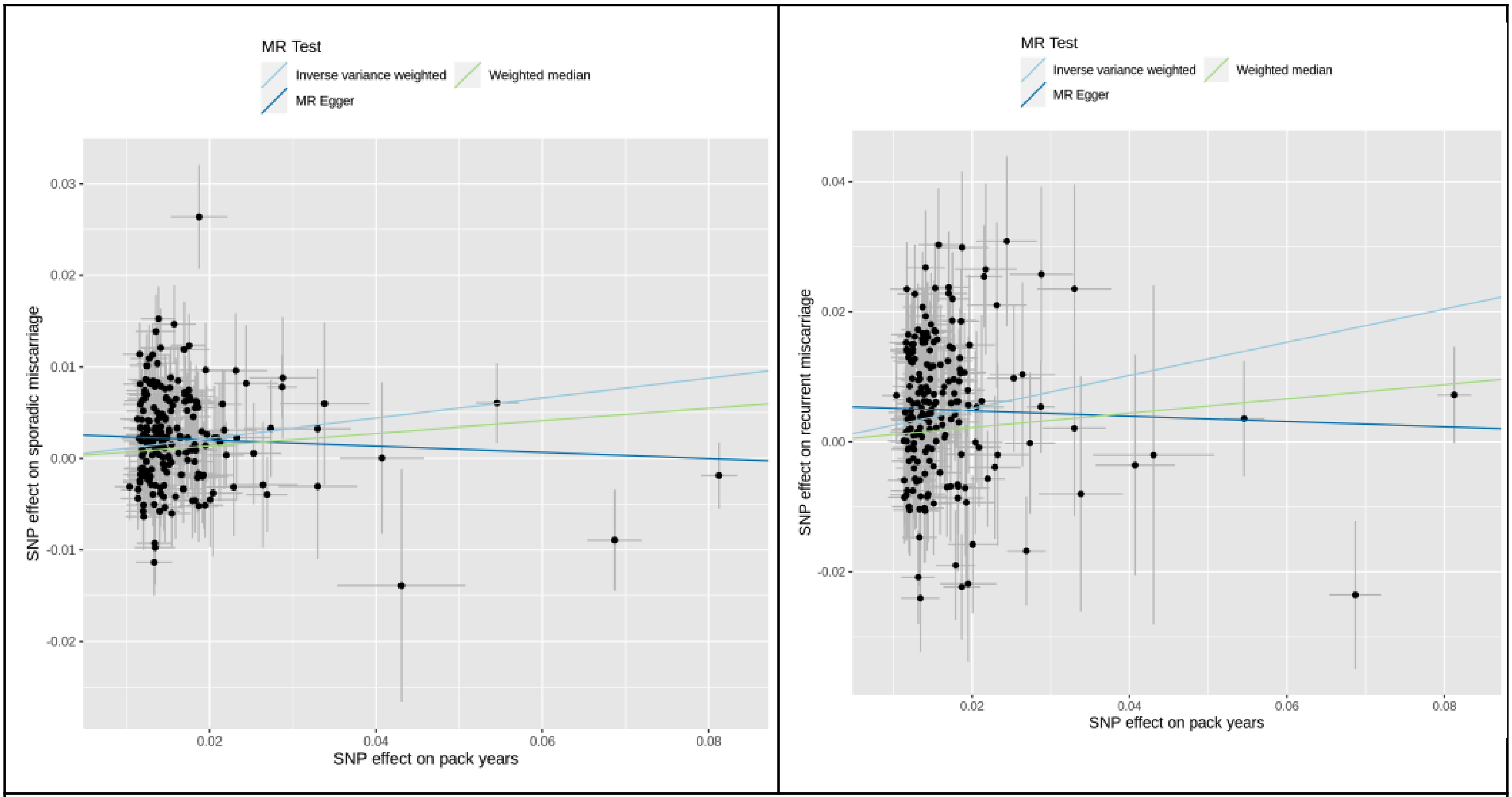


**Caffeine variables**

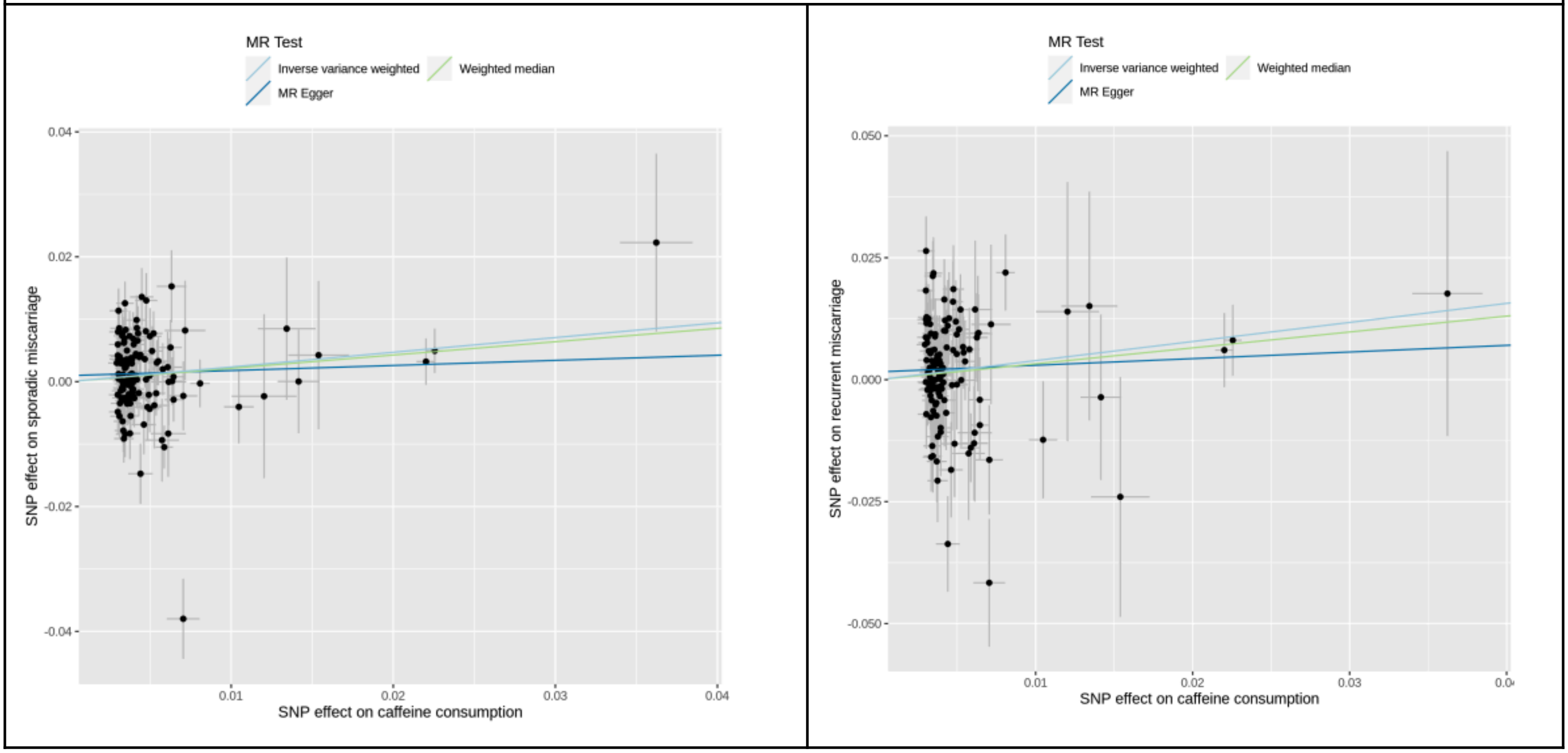

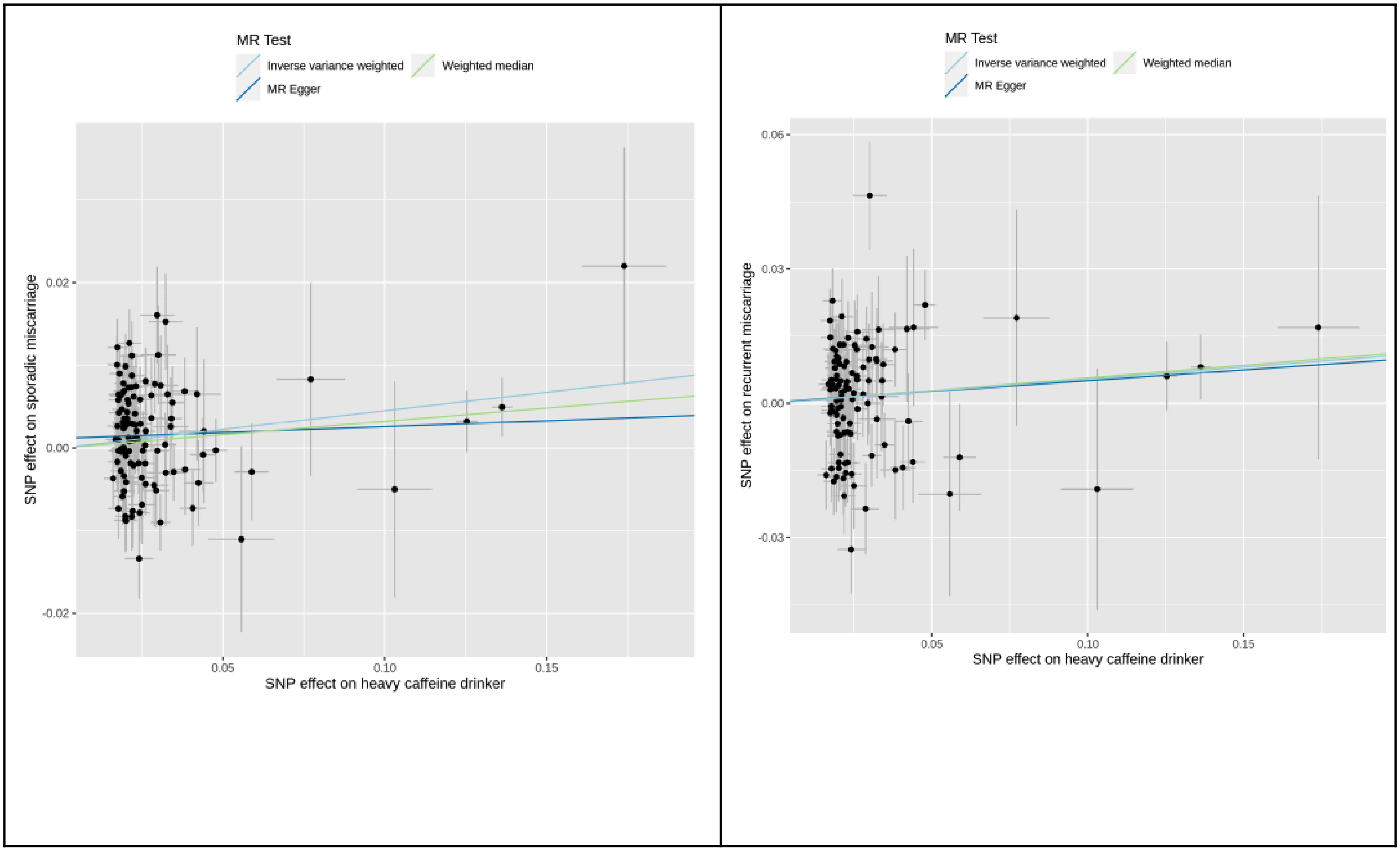

MR Test
Inverse variance weighted
Weighted median
MR Egger
SNP effect on sporadic miscarriage
SNP effect on heavy caffeine drinker
0.02
0.00
-0.02
0.05
0.10
0.15
MR Test
Inverse variance weighted
Weighted median
MR Egger
SNP effect on recurrent miscarriage
SNP effect on heavy caffeine drinker
0.06
0.03
0.00
-0.03
0.05
0.10
0.15

**Supplementary Figure 7: Split-sample Mendelian randomization estimates of smoking and caffeine intake and risk of sporadic and recurrent miscarriage**

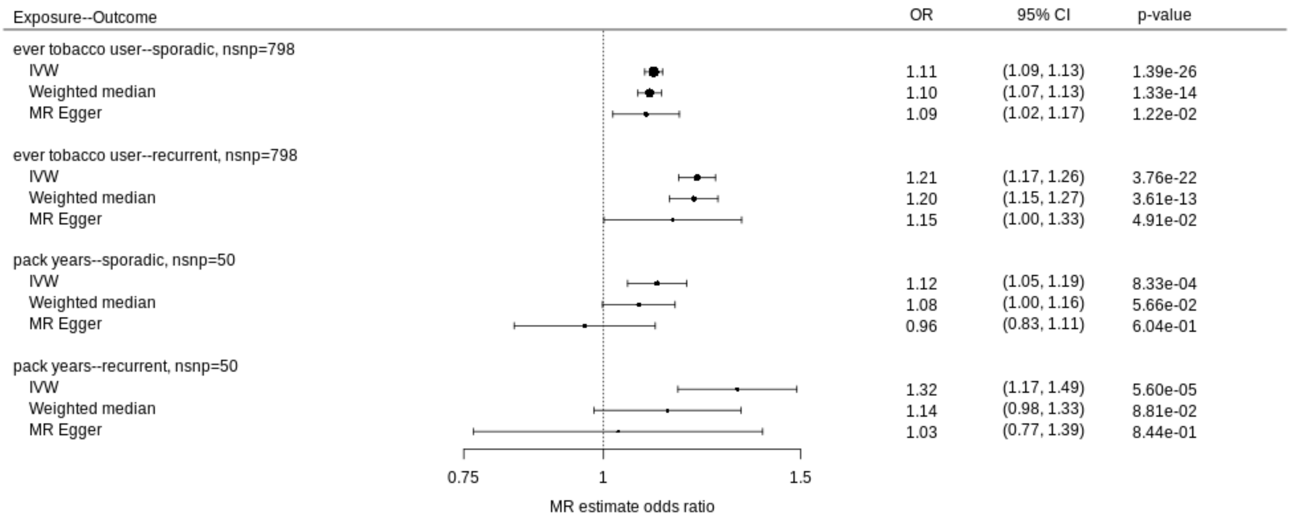


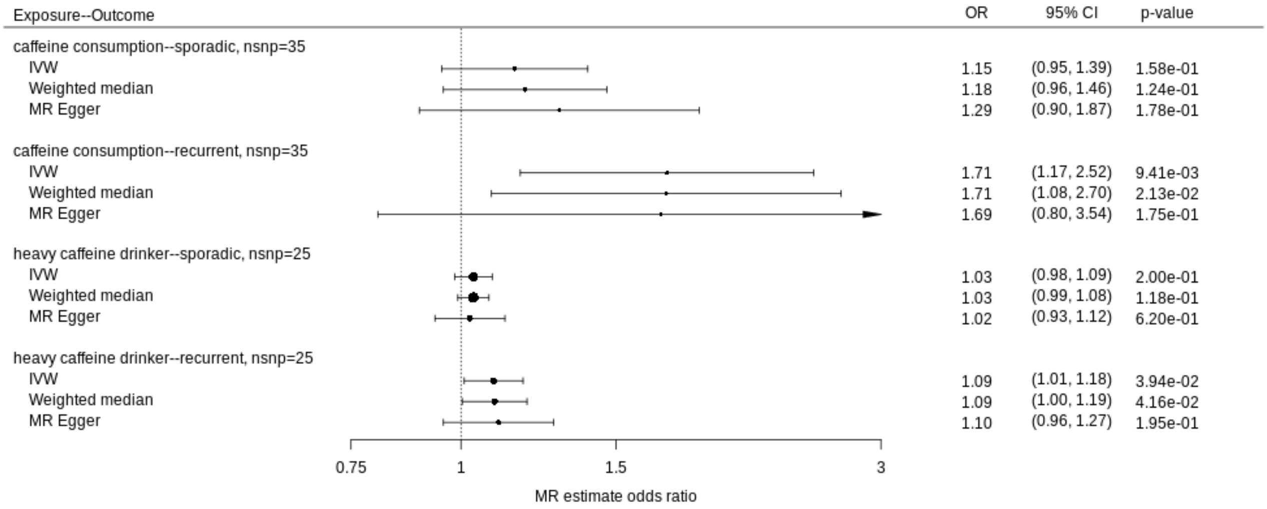

#### Supplementary Figure 8: MR Scatter plots for split-sample design

| Sporadic miscarriage | Recurrent miscarriage |
|---|---|
| **Tobacco variables** | |

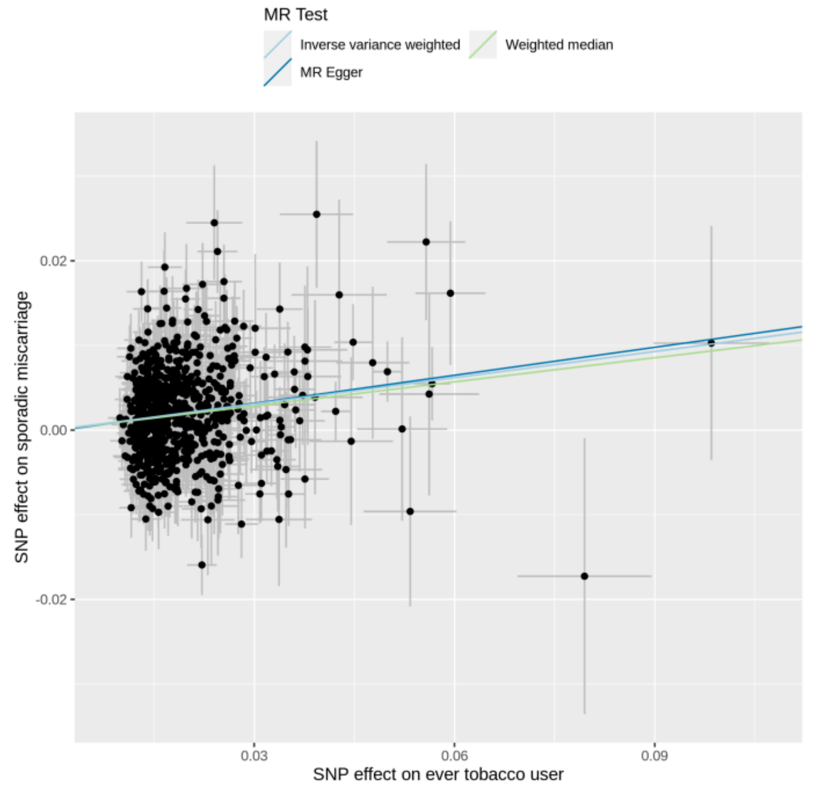


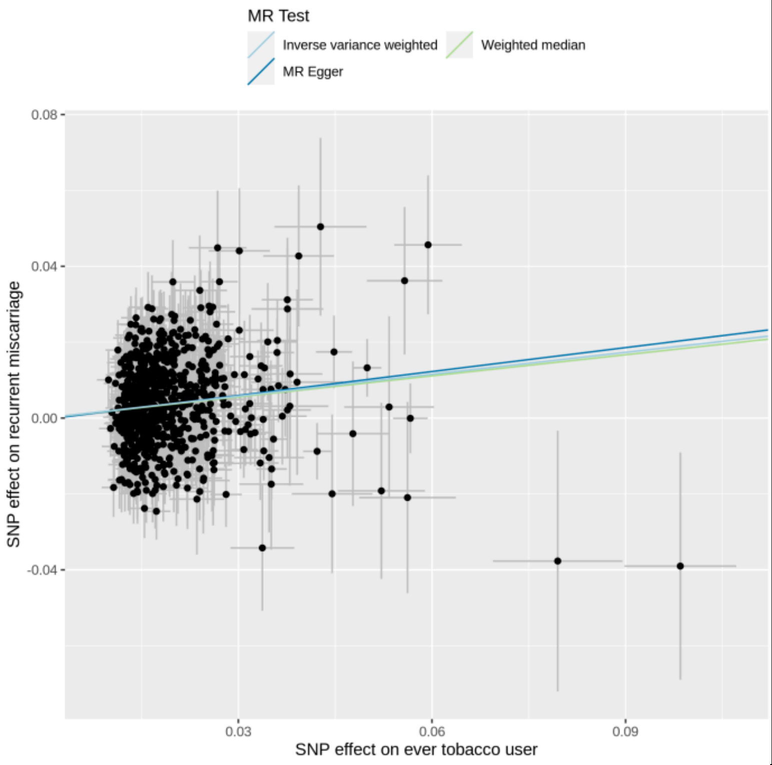

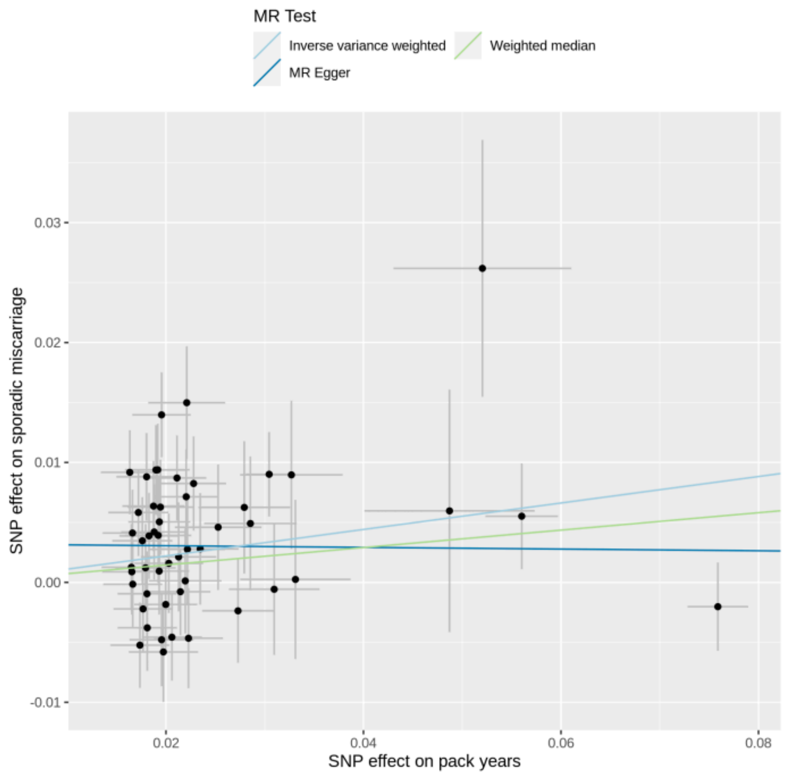


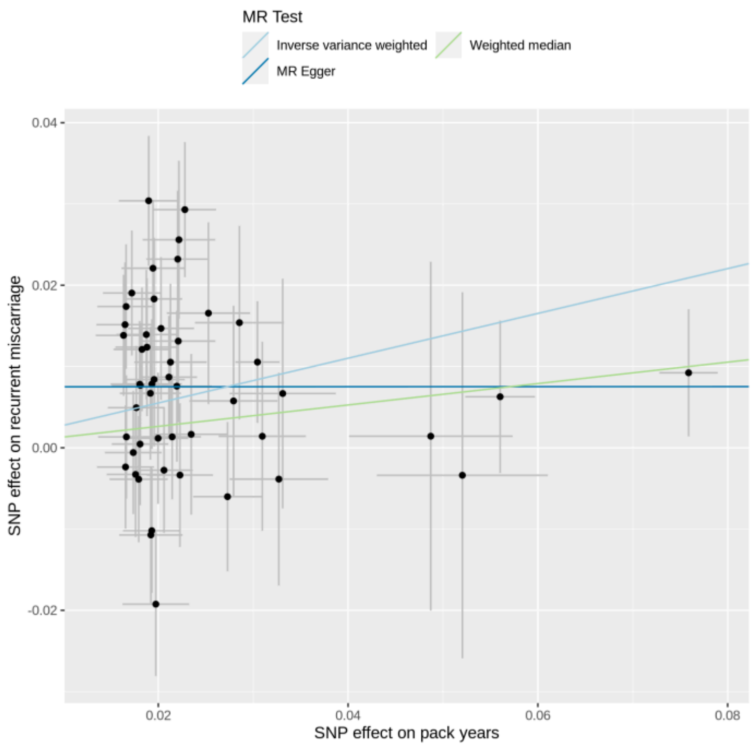


#### Caffeine variables

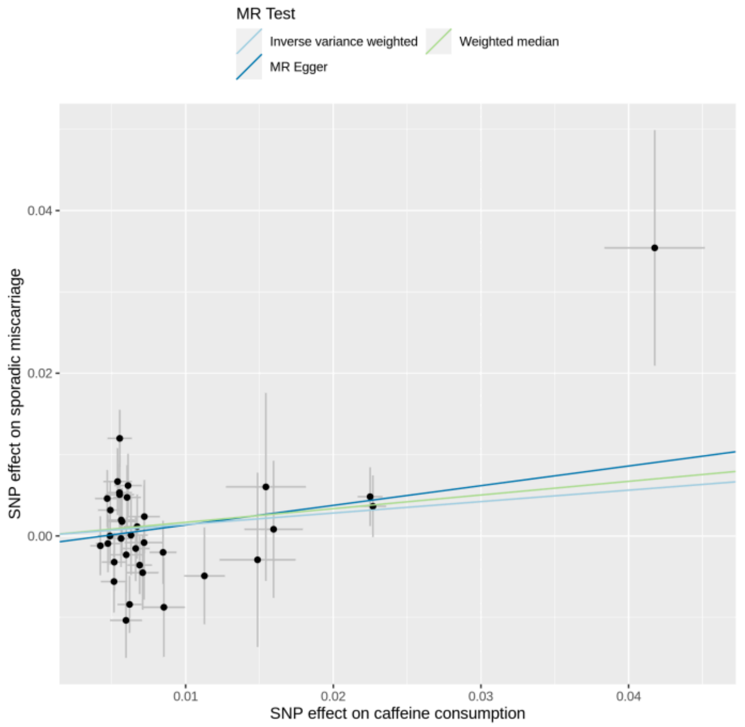


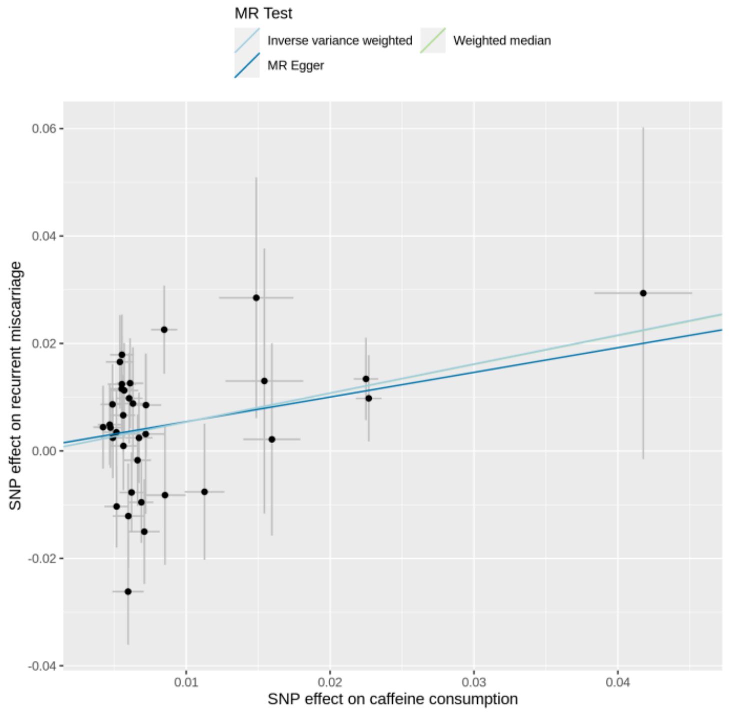

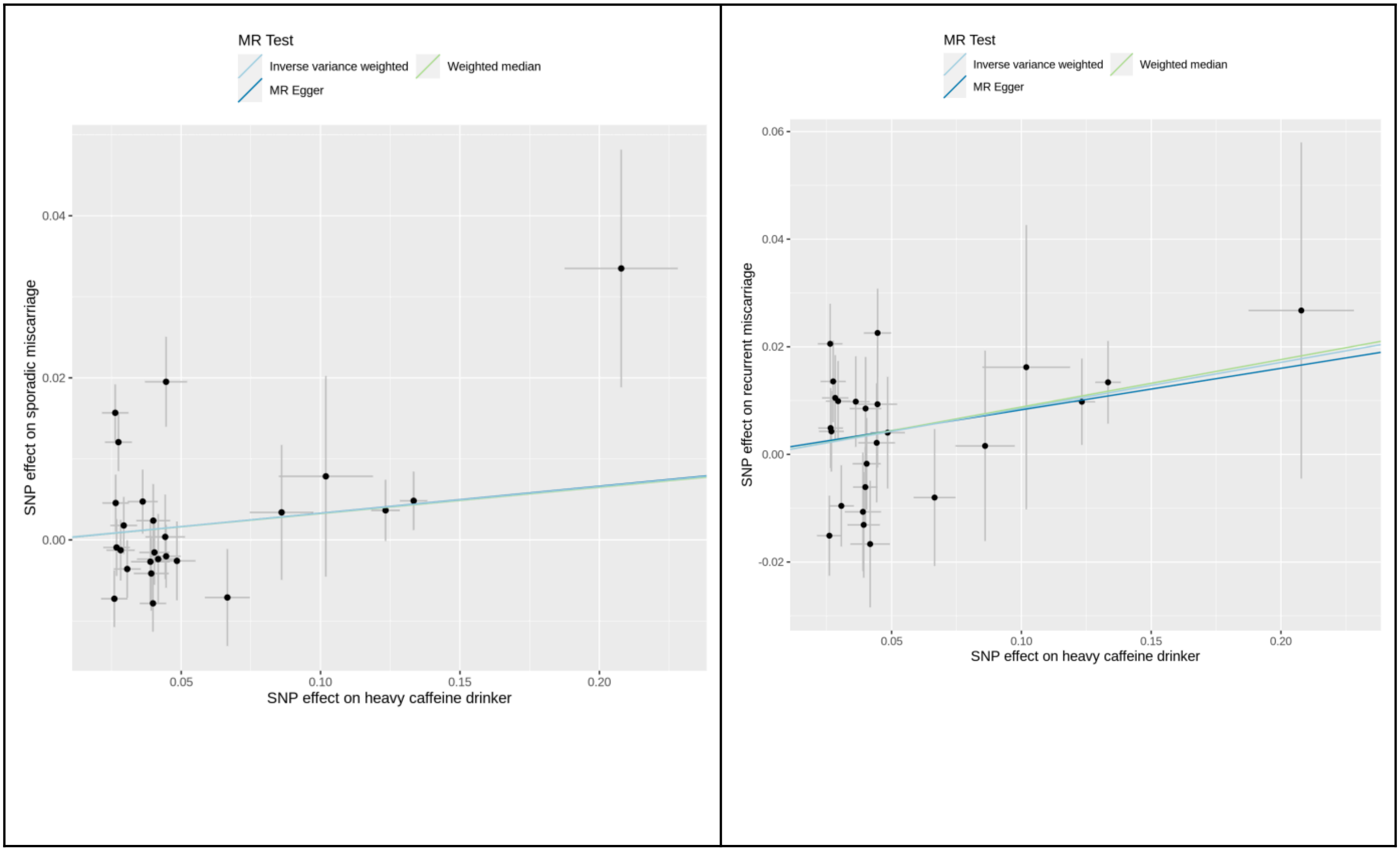
MR Test
Inverse variance weighted
Weighted median
MR Egger
SNP effect on sporadic miscarriage
SNP effect on heavy caffeine drinker
0.04
0.02
0.00
0.05
0.10
0.15
0.20
MR Test
Inverse variance weighted
Weighted median
MR Egger
SNP effect on recurrent miscarriage
SNP effect on heavy caffeine drinker
0.06
0.04
0.02
0.00
-0.02
0.05
0.10
0.15
0.20

**Supplementary Figure 9: Effect size forest plots for trans-ancestral sporadic and recurrent miscarriage top hits**

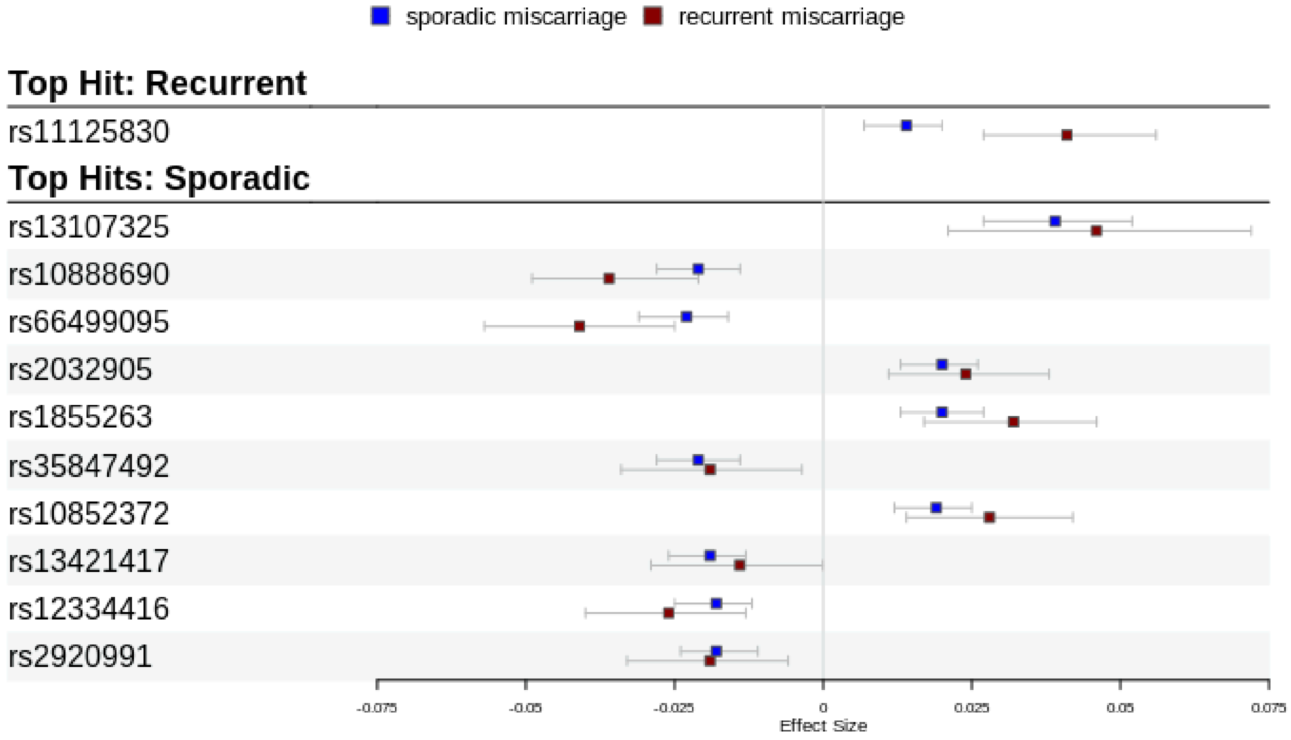
